# Non-equilibrium dynamics of the neocortex in Parkinson’s disease

**DOI:** 10.1101/2025.08.09.25333337

**Authors:** Pascal Helson, Elias Benyahiya, Sreekanth Manikandan, Josefine Waldthaler, Mikkel C. Vinding, Daniel Lundqvist, Per Svenningsson, Dhrubaditya Mitra, Arvind Kumar

## Abstract

**Background:** How dopamine depletion in Parkinson’s disease (PD) and subsequent dopamine replacement therapy (DRT) affect brain activity is poorly understood. Typically, brain activity is analysed for its spectral properties and the temporal structure of the activity is often ignored.

**Methods:** We quantified the time irreversibility of cortical activity in terms of entropy production rate (EPR). Using the Neural Estimator for Entropy Production (NEEP) algorithm on source-reconstructed resting-state magnetoencephalogram (MEG) data, we estimated the EPR in various brain regions of persons with PD (PwPD) (n = 17) and matched healthy controls (HC, n = 20). PwPD were recorded in two conditions: OFF and ON DRT (one hour after levodopa intake). HC were also recorded in two sessions separated by one hour. Motor symptoms were assessed with Movement Disorders Society-revised Unified Parkinson’s Disease Rating Scale (MDS-UPDRS-III).

**Findings:** Despite the lack of significant group differences in EPR between the HC and PD groups, we found a positive correlation between DRT induced improvement in motor symptoms (as measured by relative change in MDS-UPDRS-III scores (OFF-ON/OFF+ON)) and change in EPR. In particular, EPR changes in sensory (visual and auditory) regions and the information hub region called temporo-parieto-occipital junction were more strongly correlated with improvement in motor symptoms. Overall DRT tended to reduce EPR in PwPD and bring it close to the EPR values of HCs. Furthermore, PwPD with a higher EPR than HC showed better response to medication correlated with an increased EPR.

**Interpretations:** Higher EPR in PwPD than HCs suggest that chronic dopamine loss alters local network interactions within cortical regions so as to reduces diversity of brain state transitions. Our analysis also opens a promising avenue to extract medication effects from non-non-invasively acquired cortical activity.

**Funding:** Digital Futures, StratNeuro, NatMEG experiments were funded by Stiftelsen for Strategisk Forskning, DM acknowledges the support of the Swedish Research Council Grant No. 638-2013-9243.

## Introduction

Progressive and major loss of dopamine is a cardinal feature of Parkinson’s disease (PD). The consequences of this loss on the neural circuits in the basal ganglia (BG) and motor regions are fairly well characterised.^1^ However, much less is known about reorganisation of cortical activity due to the loss of dopamine and other PD related changes. This understanding is necessary to develop clinically useful biomarkers of PD and to quantify the impact of various therapies based non-invasive brain activity.

Typically, non-invasively measured brain activity such as EEG/MEG are analysed for their spectral content (e.g. power in different frequency bands) or functional connectivity.^2,3^ Such methods ignore the temporal structure of brain activity, therefore, we lack information on how dopamine loss affect the temporal dynamics of brain activity. Here, we introduce a new way of analysing EEG/MEG which is based on the fundamental non-equilibrium nature of brain dynamics. Using this approach on MEG data, we demonstrate how DRT may help restore cortical brain dynamics in PwPD.

EEG/MEG measures collective activity of neuronal networks in the brain. Given the noise in the latter and sensory inputs, these networks are best described as stochastic dynamical systems.^4,5^ The activity generated by such stochastic dynamical systems could be stationary or non-stationary (in statistical sense) and in thermal equilibrium or non-equilibrium. The analysis of brain activity shows that, at least in controlled experimental conditions, the brain activity is in a stationary state. This assumption justifies various estimates of the statistics of brain activity. By contrast, it is not common to test whether the brain dynamics are in equilibrium or non-equilibrium. As Schroedinger argued,^6^ living systems must operate away from their equilibrium. The same is true for the brain as an information processing dynamics system. The brain activity at some spatio-temporal scales should be pushed out of equilibrium as it dynamically represents the environment by mixing external inputs with internal state and memory. It is not well understood to what extent the brain dynamics is operating far from equilibrium and how PD related changes affect this non-equilibrium state at the macroscopic spatial scale measured by MEG signals.

A key feature of non-equilibrium state is that at few specific spatial and temporal scales, time series generated by the system are not time reversible. Hence, non-equilibrium introduces the so called ‘arrow of time’. Time irreversibility of a time series is closely linked to ‘entropy production rate’ (EPR).^7^ Recent advances in stochastic thermodynamics allow us to calculate a lower bound on EPR from a time series without explicitly knowing the equations governing the dynamics of the system.^8^ We exploit this approach to estimate EPR using MEG recordings from healthy controls (HC) and PwPD (OFF and ON medication, meaning after 12-hour medication withdrawal from all dopaminergic medication and one hour after levodopa intake, respectively).

Our results show that in general, most of the brain regions – except in the left dorsolateral prefrontal cortex and the orbital and polar frontal cortex – have a higher EPR in PwPD compared to HC. However, EPR showed large variability both across brain regions and across individuals, reducing the statistical power of PwPD-HC group comparisons. Next we found that DRT tended to reduce the EPR, especially in PwPD who showed large EPR in OFF-medication state. More importantly, we observed a levodopa-induced improvement in motor function (decrease in the Unified Parkinson’s Disease Rating Scale: UPDRS-III)) was correlated with a decrease of EPR values. This correlation was strongest in the temporo-parieto-occipital junction of the right hemisphere (TPOJ-rh). Interestingly, the same region in the left hemisphere was the one with the most increased EPR in the PwPD-HC comparison, hence highlighting the important role of this brain region in PD. Finally, breaking down the UPDRS into the main motor symptoms revealed a positive correlation between change in EPR and change in bradykinesia scores.

**Research in context**

**Evidence before this study**

Spectral properties (e.g. beta band oscillations, aperiodic activity) of the population activity of the brain are altered in patients with Parkinson’s disease (PD). But it is not known how the temporal structure of such activity is changed in Parkinson’s disease and levodopa medication.

**Added value of this study**

Here we put forward a new approach of electrophysiological signal analysis in the context of MEG of persons with PD. It goes beyond classical spectral analysis using a physics-grounded property of the signal called entropy production rate (EPR). Aligned with the idea of brain structure to brain dynamics alterations, our findings on entropy production rate highlights changes of the signal’s irreversibility in PD. These changes specifically correlate positively with the effect of the treatment, inviting to consider EPR as a treatment effect biomarker. In addition, it shows the interest and necessity of individual/longitudinal analysis in the complex system framework of brain signals: brain dynamics subject-wise differences hinder the use of population analysis.

**Implications of all the available evidence**

We provide evidence that non-invasive signals such as MEG can provide information about the effect of dopamine replacement therapy and give us new insights into how cortical dynamics is changes due to PD related changes.

## Methods

### Data

#### Data acquisition

We use the same source-space resting state data as in the work by Helson et al.^9^ which are part of the data recorded for the study by Vinding et al.^10^ Data were recorded on 37 age-matched non-demented participants, 17 PwPD and 20 HC, all of them for two sessions (more details in Table <u>T1</u>). PwPD were recorded OFF medication (≥ 12 hours after DRT) and ON medication (1 hour post-dose), while HCs followed the same protocol without medication. MEG recordings were done at the Swedish National Facility for MEG <u>(NatMEG)</u> using the Elekta Neuromag TRIUX 306-channel MEG system. Individual structural MRI scans were collected for the source reconstruction.

#### Data preprocessing

More details on the source reconstruction methods used for these data can be found in previous works.^10,9^ We here used the resulting time series of the magnetic field measured at 8196 sources (4098 per hemisphere) covering the whole cortical surface. Using the function *mne*.*extract label time course* with *mode*=*None* in MNE Python,^11^ we labelled the sources with the HCP-MM1-combined atlas^12^ (44 brain regions). Unlike standard methods that average voxel signals by region, we retained vertex-wise dynamics to preserve information, as EPR analysis tolerates correlated time series. The MEG data were downsampled from 1000Hz to 200Hz, and then low pass filtered at 45Hz to avoid artefacts such as muscle and power-line artefacts.

#### Ethics

Due to privacy concerns regarding the PwPD’s identities, the data is exclusively accessible in Stockholm and only by personnel verified by the NatMEG. We received approval for this study from the local ethics committee (Etikprövningsnämden, dnr 2016/911-31/1, last updated with amendment dnr 2024-00658-02). Written and oral informed consent from the participants was also obtained before their participation following the Declaration of Helsinki.

### Entropy production rate inference

We estimated the EPR of the MEG time series’ trajectories using the framework of stochastic thermodynamics. EPR can be interpreted as a quantification of the irreversibility of a process.^13^ When a stochastic process (**x**(*t*))_*t ≥* 0_ is in a non-equilibrium steady state (NESS), the thermodynamic uncertainty relation (TUR)^14^ establishes a lower bound on its EPR, expressed as:

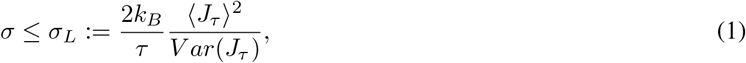

where *k*_*B*_ is the Boltzmann constant, *J*_*T*_ is any current of (**x**(*t*))_*t ≥*0_ integrated during an observation time *T, σ* is the EPR, and ⟨·⟩ and *V ar*(·) stand for respectively the statistical mean and variance. To infer the EPR, we use a method consisting in maximising this lower bound with respect to the current and thus we obtained the optimisation problem:^8^

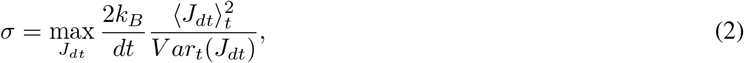

Where 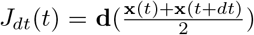. (x(*t* + *dt*)-x(*t*)) is a scalar and **d**(x) a vector. We used a neural network model (Fig. 1C) to find the function *d* that maximises *ω* with *J*_*dt*_. We provide a general intuition on this property in the EPR part of the supplementary information.

**Figure 1.**
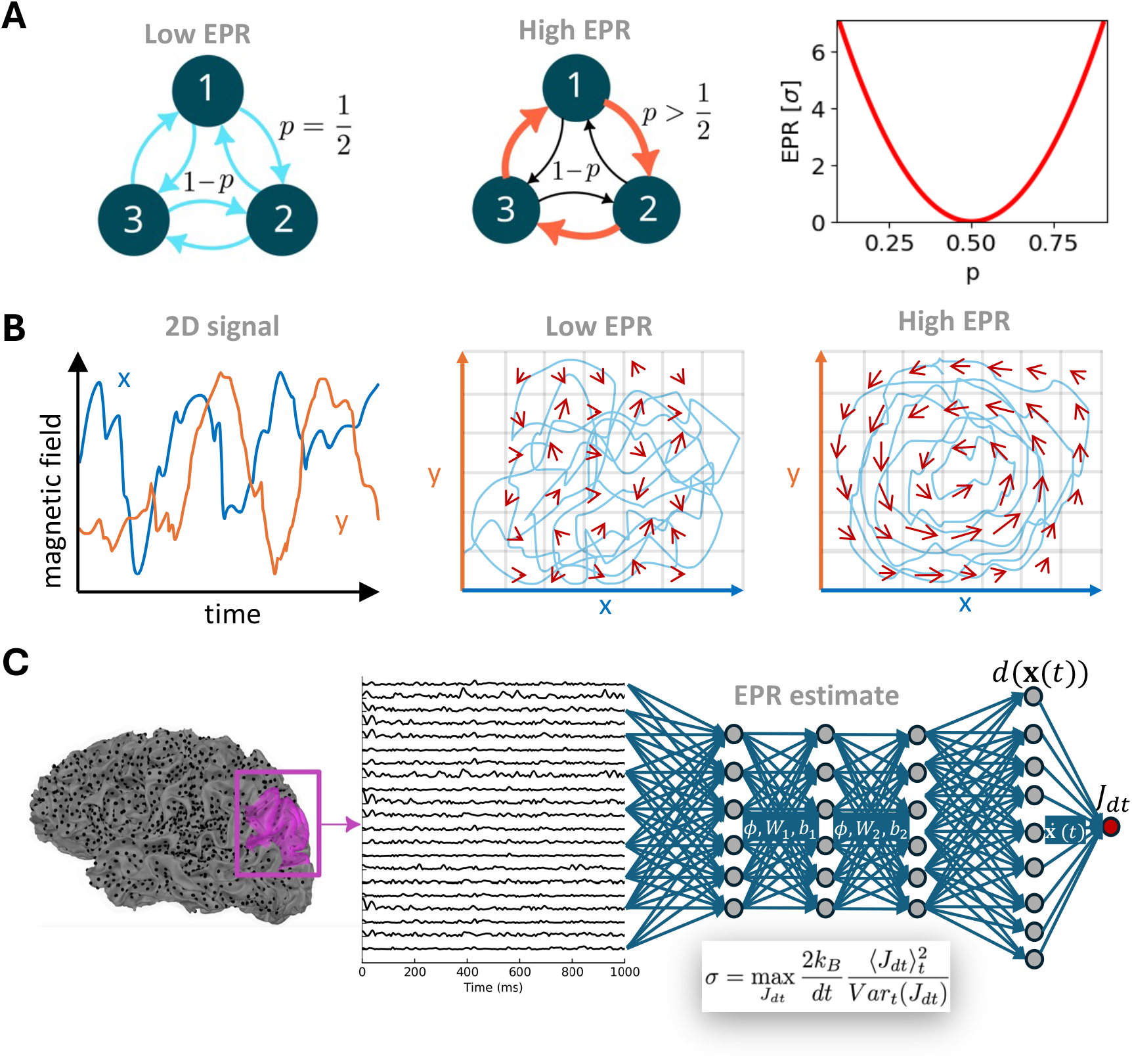
Entropy production rate and its inference from MEG data. **(A)** Diagram of a three state system. The state transition probabilities are illustrated with the colour of the arrows. Left: Detailed balance case when all transition rates are equal, so the net current is null; EPR is zero. Middle: Broken detailed balance case: for 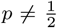, the transition rates between two states are not equal and the system reaches a non equilibrium steady state (NESS), with a non zero constant net current. This corresponds to strictly positive EPR. Right: EPR as a function of the state transition probability *p*, derived from analytical computations. **(B)** EPR in continuous 2D signals. An example of such signal is given in the left panel. In the middle and right panel, a similar signal is plotted in the phase space respectively for a low and high EPR case. The red arrows indicate the current fluxes, estimated as the average transition probability from one grid cell to another. Longer and more aligned arrows correspond to a larger EPR. **(C)** Overview of the EPR estimation pipeline for the MEG data. Left: Illustration of sources’ locations on a sample brain. EPR was calculate for well defined brain regions. One example region is highlighted in purple. Middle: Time series of sources from the purple region. Right: Schematic of how the function *d* – see equation (2) – is found using a CNN to get the maximal EPR σ from the MEG time series.

We clustered the data into 44 regions (containing ≈ 70 to 370 sources) and computed one EPR value per region by feeding each region’s source time series into the neural network. Because the number of sources in each region was different, we checked that such disparity did not bias the EPR estimation (Fig. S1).

### Statistical analysis

#### Data cleaning

We remove one PD patient because the second session recording was too short compared to the other subjects (less than 2 minutes compared to 8 minutes).

To validate the MEG EPR estimates, we compared them to surrogate data with randomised phase, rejecting subjects whose EPRs were not significantly different (Wilcoxon test with p-value *<* 0.001, Fig. S2); this removed one HC and confirmed that the following effects arise from temporal, not just spectral, structure.

Brain regions were excluded if their original and surrogate EPRs were not significantly different in any group (p-value*>* 0.001), removing one region (Orbital and Polar Frontal Cortex right hemisphere, see Fig. S3), or if they lacked test-retest reliability across HC sessions (*p* – *value >* 0.05, FDR-corrected), which removed none (Fig. S4).

#### Comparing the EPRs between groups

After filtering unreliable data, we used permutation MANOVA (using `permanova` function of the Python package *scikit-bio* with 200 000 permutations, p-value*<* 0.05) to compare HC and PD groups (Fig. 2A), followed by region-wise Mann-Whitney tests with FDR correction using permutation-based Benjamini-Hochberg adjustment (Fig. S5).

**Figure 2.**
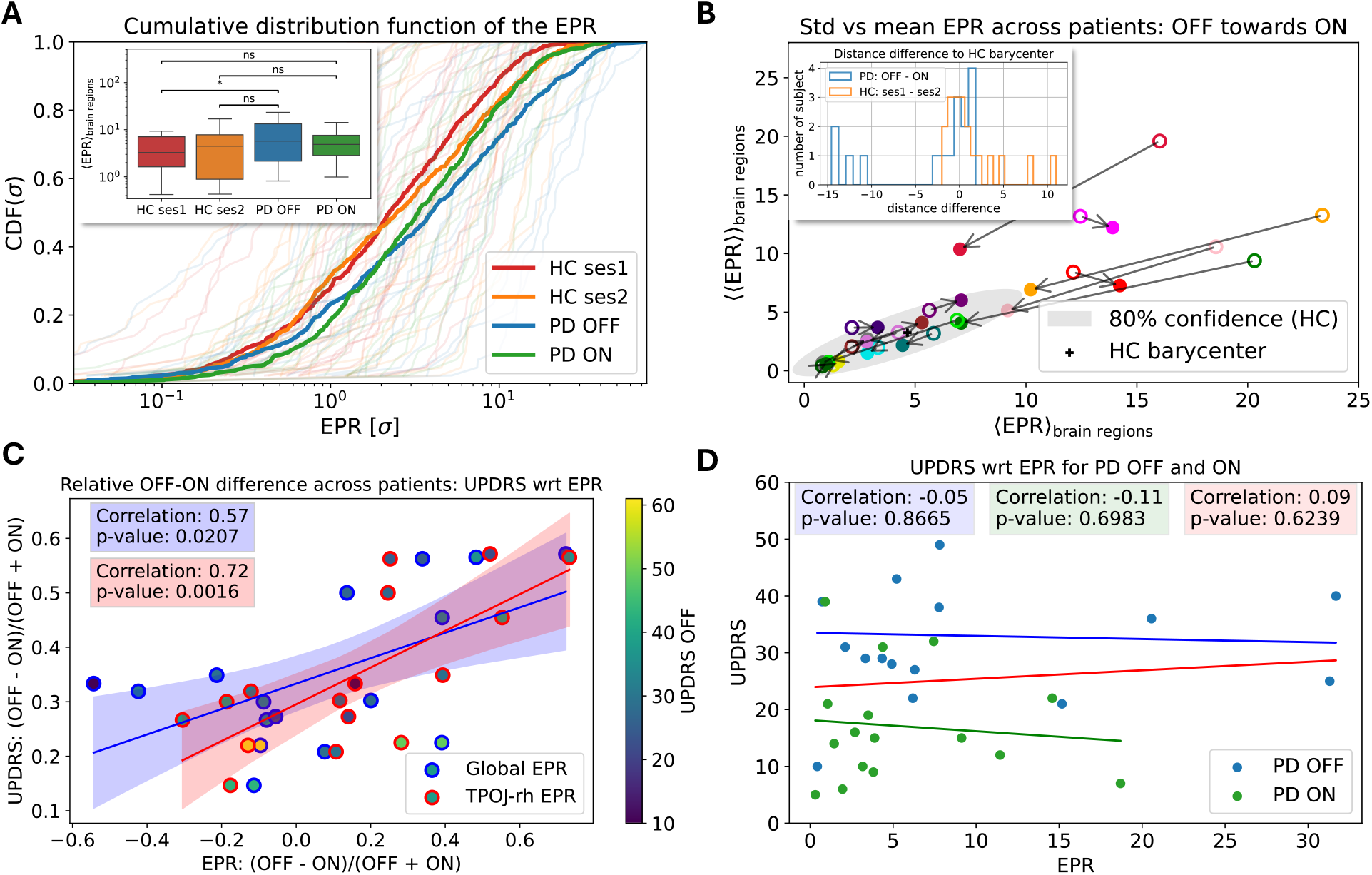
EPR changes between groups and with respect to medication in the PD groups. **(A)** Cumulative distribution function for each subject in light colours and their group counter part in bold. The inset shows the results of permutation MANOVA statistical tests between the groups. The values that compose the box plots are averaged over brain regions. **(B)** DRT induced change in the EPR statistics (mean and SD). The grey zone indicates where most of the HC (mean, SD) dots are which is an ellipse centred in the black cross. Each arrow corresponds to the EPR statistics of a subject. Arrows start at OFF-medication state (empty dots) and the arrowheads indicate ON-medication state (filled dots). **(C)** DRT induced modulation of the EPR mean over brain regions (blue border) – or the EPR of the TPOJ-rh region (red border) – and UPDRS motor tests. Each dot indicates the EPR and UPDRS score of one subject. The solid lines are best linear fits. The shaded area represents the 95% confidence interval of the regression line. **(D)** Correlations and the best linear fits (solid lines) between EPR mean and UPDRS in ON (green), OFF (blue) and both (red) medication states. Each dot is one subject and one session.

#### Comparing the EPRs within groups over sessions

We analysed session-wise EPR mean and SD changes in HC and PwPD. Defining a healthy area (covering 80% of HC data) using PCA and Gaussian-based ellipses, we then measured session changes via Euclidean distance to the HC ellipse centre (Fig. 2B and S8).

#### Testing the EPR link with UPDRS

We used the part III of the Movement Disorders Society-revised Unified Parkinson’s Disease Rating Scale (MDS-UPDRS-III) to assess the motor symptoms. In the following, we will use the acronym UPDRS. Unless otherwise specified, UPDRS will refer to the total MDS-UPDRS-III score.

We need a definition to clarify what follows: we define the relative (with respect to medication) value *R*(*X*) of a variable *X* as

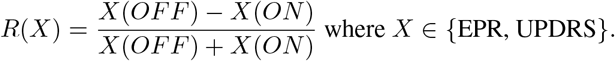

In other words, we calculated the medication-induced change in these measures relative to the sum of their OFF and ON values. We performed a linear regression between the relative EPR and UPDRS. The advantage of the relative quantity is its removal of the linear dependence of all variables not depending on medication (like age for example). We sub-divided the UPDRS into core motor symptoms gathering the total UPDRS scores as described by Goetz et al.^15^ into its posture = {F1}, bradykinesia = {F4, F5, F7}, tremor = {F2, F6}, and rigidity = {F3} components.

The regression coefficients and p-values were obtained using the `linregress` function of the *scipy*.*stats* package on Python, see Fig. 2C,D. The latter uses a two-sided Wald test with t-distribution of the test statistic where the null hypothesis is that the slope of the regression line is zero.

## Results

### The idea of entropy production rate

To introduce the idea of entropy production rate, let us first consider a simple systems with three states (Fig. 1A, left). Here, the system makes transition between the different states with equal probability, *p* = 1*/*2. In this system, all the states are equally probable and the rate of transition between them are equal, by design. Thus, a time series generated from this system is (e.g. 1 → 2, 2 → 3, 3 → 1) is statistically indistinguishable from its time reversed version (1 → 3, 3 → 2, 2 → 1). Such a dynamics is a signature of thermal equilibrium and the system’s EPR is zero.

However, if *p* ≠ 1*/*2, i.e. there is an asymmetry in the system, some state transitions are more likely than others. For example, in the toy model depicted in the middle of Fig. 1A, 1 → 2, 2 → 3, 3 → 1 is more likely than its time reversed version. Such systems are said to be operating away from the thermal equilibrium and have a positive EPR. In this example, by varying *p* we can push the system out of equilibrium such that it has positive EPR (Fig. 1A, right). Here a non-zero EPR emerges as a result of asymmetry in the system.

For neural activity, there are no discrete states as we considered in our toy example (Fig. 1B, left). For such analogue systems, we can discretise the activity space (Fig. 1B, middle&right) and estimate the average direction in which the activity moves out of a particular voxel (Fig. 1B, middle&right, red arrows). In this context, a system in equilibrium has no consistent flow (Fig. 1B, middle), whereas a system operating in its non-equilibrium regime will have a clear flow direction (Fig. 1B, right). This idea of discretised states is very intuitive and EPR of such systems can be approximated numerically as explained. However, it is not easy to extend such analysis to high dimensional continuous data such as MEG. This is why we used a neural network to optimise the lower bound derived from the Thermodynamic Uncertainty Relation (TUR) inequality (eq (2) and Fig. 1C).

### EPR distributions remain similar at the group level

We measured resting state MEG for 44 brain regions as defined by the HCP-MM1 combined atlas.^12^ We used these time series to estimate the EPR of each brain region using a method similar to the Neural Estimator for Entropy Production (NEEP) algorithm.^16^ The question that we address in this work is how far from equilibrium are the brain regions operating in PD and under DRT.

The EPRs across the brain have a spatial structure with small EPR in the frontal regions to high EPR in the occipital regions both in HC and PD groups (Fig. S6, left). However, the variability across subjects was higher in the central regions of the cortex, especially in the PD group (Fig. S6, right). More details are given in the supplementary tables recapitulating the EPR for each subject and brain region (Fig. T2T3T4T5).

To compare different subjects and groups, we constructed cumulative distribution functions (CDF) based on all 44 EPR values for each subject (light lines in Fig. 2A). There was high variability in the distribution of EPRs in the subjects, both HC and PwPD (Fig. 2A and its box plot inset). The mean of the EPR distributions (thick lines in Fig. 2A) remained similar between the groups. Nevertheless, there was a significant increase in EPR between HC in the first session and PD OFF (see the box plot inset in Fig. 2A, p-value 0.02). This difference vanished when we analysed the second session for HC. Interestingly, we observed a decrease both in the median and the SD of the mean – over brain regions – EPR values between PD OFF and PD ON (compare steepness of the blue and green CDF in Fig. 2A and the box plot inset). Conversely, the SD increased between sessions 1 and 2 for HC (compare steepness of the red and orange CDF in Fig. 2A and the box plot inset). Overall, PwPD showed a tendency toward higher EPR than HC, though not statistically significant. Under DRT, PwPD exhibited a narrower EPR distribution (lower SD) and reduced median, whereas the second session of HC displays greater median and variability (higher SD).

We also compared the EPRs for each brain region separately but no EPR difference (between HC and PwPD) crossed the significance threshold when we corrected for multiple comparisons (FDR). Still it is worth pointing that the temporo-parieto-occipital junction (TPOJ) and the early auditory cortex (EAC), both in the left hemisphere, showed notable increase of the EPR in PD OFF as compared to HC groups (see Fig. S7A,C).

### EPR spatial statistics improved by medication

Because the changes in the EPR distribution over sessions varied in opposite directions depending on the group (Fig. 2A), we checked how they changed across the two sessions (HC ses1 vs ses2 and PD-OFF vs PD ON) for each subject. Although there was high individual variability, healthy controls showed only small changes in the EPR mean and SD across the two sessions (Fig. S8). We thus defined an ellipse based on the two first components of the PCA of the HC’s EPRs. The latter contains more than 80% of such data as can be seen in Fig. S8. In PwPD, DRT led to larger changes in the EPR mean and SD (arrow lengths in Fig. 2B). Interestingly, larger EPR mean and SD were consistently reduced by a large amount. Thus, there was a tendency that DRT pushed the brain activity of PwPD towards healthy levels of EPR (Fig. 2B, see also Fig. S8 and S9). This can be seen in the inset of Fig. 2B showing the distribution of the differences in this distance between sessions, specifically session 1 minus session 2. Indeed, when there was a large change of the distance to the HC barycentre, it was always towards the HC barycentre (negative values in the histogram). On the contrary, HC tended to have higher EPR mean and variance on the second session. Such distance makes sense as both the SD and mean have the same unit. This distance captures differences with the healthy group both in terms of absolute value of the mean and heterogeneity between brain regions. Screening over brain regions showed that some brain regions (like the EAC-lh) were more affected (Fig. S7D).

### Changes in EPR are correlated with DRT induced changes in UPDRS

Next, we found a positive correlation between the relative changes of EPR and relative changes of UPDRS (*P* = 0.57, p-value = 0.02, Fig. 2C). Interestingly, this correlation was even stronger when we considered the change in EPR specifically in the right temporo-parieto-occipital junction (*ρ* = 0.73, p-value = 0.0016, Fig. 2C) and the early right auditory cortex (*ρ* = 0.59, p-value = 0.017). It is important to note that these latter analyses were conducted post-hoc based on observed group findings and did not include corrections for multiple comparisons.

Next, when grouping the UPDRS-III sub-scores into broader symptoms, we only obtained a significant correlation with bradykinesia (*ρ* = 0.54, p-value = 0.03, Fig. S10 and *ρ* = 0.71, p-value = 0.002 in TPOJ-rh region).

Given such strong correlations between relative changes of EPR and UPDRS, we expected a similar relationship between EPR changes and absolute UPDRS. However, EPR or EPR changes were not correlated with UPDRS (Fig. 2D). In addition, we did not find any correlation (with both original values and relative ones) with other variables such as age, disease duration, levodopa equivalent daily dose (LEDD), and scores of depression, anxiety (HADS) and cognition (MoCA) (see Fig. S11).

## Discussion

### EPR of MEG as a possible readout of medication success

Here, we observed that together with the amelioration of motor symptoms, the statistics of the EPR also tended to return to normal levels after medication. These results suggested a possible link between the EPR and the motor performance. Interestingly, rather than a direct EPR-to-UPDRS correlation, we found a positive correlation between relative changes in UPDRS and relative changes in EPR: when the effect of medication was stronger, the EPR decreased and vice and versa. In other words, the patients who showed a decrease in EPR post-medication were also the ones with the strongest clinical improvement, suggesting that the EPR reduction aligns with restored motor function at the individual level. In addition, the lack of direct correlation between the EPR and UPDRS highlights the link of the EPR to the effectiveness of the medication on motor symptoms rather than the UPDRS scores alone. Finally, bradykinesia was the only motor symptom with a significant correlation (Fig. S10). Bradykinesia is the core PD motor feature that has also shown the most consistent relationship with other oscillatory and non-oscillatory neurophysiological measures across subcortical and cortical brain areas. Although some correlations were found previously between bradykinesia scores and STN beta oscillation or sensorimotor-cortical beta^17,18^ or beta burst rate,^10,19^ the DRT effects on the latter remain unclear.^20^ Our findings highlight the potential of EPR analysis in addressing this question.

Overall DRT brought the MEG activity to more normal EPR statistics. However, the distribution of relative EPR among PwPD suggests two patient subgroups (Fig. 2B,C): those sensitive to medication (positive relative EPR) and those resistant to medication (negative relative EPR). There is no clear pattern of the UPDRS among these two groups (Fig. 2C). Adding the fact that the EPR is not correlated with UPDRS (Fig. 2D), this suggests that the network mechanisms underlying PD are different for PwPD with high and low EPR and underscores EPR’s potential as a treatment biomarker of network dysfunction.

### The role of the temporo-parieto-occipital junction and sensory areas

Region-wise comparisons of EPR revealed the importance of two brain regions (with hemispheric asymmetries, see below): the temporo-parieto-occipital junction (TPOJ) and the early auditory cortex (EAC). The left TPOJ and EAC showed the most pronounced EPR increases in PwPD compared to HC. In contrast, the right hemisphere’s relative EPR of these regions positively correlated with UPDRS scores. This hemispheric dissociation aligns with the right TPOJ’s a priori role in integrating whole-body sensory feedback for movement control, while the left TPOJ changes may represent compensatory linguistic or cognitive adaptations.^21^ It is also interesting to note that these regions are also rich in dopamine receptor expression.^22^ These results also highlight that the impairment of the left hemisphere is not corrected by the DRT as systematically as its right counterpart which seems to be a better leverage against motor symptoms.

In general, most of the brain regions – except in the left dorsolateral prefrontal cortex and the orbital and polar frontal cortex – showed higher EPR in PwPD compared to HC. The parietal, and to some extent the temporal and occipital cortical regions were the most changed (see Fig. S6). Even though these differences were not significant in any region after correction for multiple comparison, they point to a reorganisation of cortical dynamics beyond the primary sensorimotor region in PD. Such consistency of higher EPR in PwPD as compared to HC suggests that sensory processing in PwPD – particularly in the visual cortex and associative parietal regions – may operate with reduced reversibility compared to HC.

### A counter intuitive increase of EPR in PwPD and neural mechanism that may control EPR

PD is associated with widespread loss of dopamine and concomitant synaptic connectivity loss.^23,1^ Intuitively, we might think that such changes should reduce EPR. Indeed, there are suggestions of lower EPR in persons with Alzheimer’s disease which is characterised by a global loss of connectivity.^24^ We here report an *increase* in EPR in PwPD and a *reduction* in EPR after DRT. This suggests that PD does not manifest in cortex as a simple loss of connectivity. Instead, our work shows that in PD, the local and global cortical dynamics are more constrained and structured which make the time series of MEG less time-irreversible.

Networks in the brain are stochastic dynamics systems and a change in the EPR could be due to a change in input, network connectivity and network dynamics. At the simplest, a change in bidirectional connectivity among the nodes of a network can alter EPR (as we demonstrated in the Fig. 1A). However, even when a system has bidirectional connections (symmetric connectivity matrix), networks can show high EPR if their nodes are driven by inhomogeneous inputs. Thus, higher EPR indicates changes in the local as well as non-local connectivity. One potential way to alter these connectivities is to alter the balance of excitation and inhibition (EI) and there is some support of altered EI balance in neocortical activity in PwPD.^9,25^ EI-balance can further influence EPR by introducing pathological oscillations and coherence.^26^ Such frequency- and region-specific changes might alter EPR for instance by enhancing directed interactions leading to beta bursts.^27,28,10^

### EPR sweet spot

Spatio-temporal sequences of neural activity are a hallmark of meaningful behaviour. Generating such activity patterns requires that the brain dynamics is pushed out its equilibrium (random asynchronous activity). EPR is a way to quantify this deviation from equilibrium. It is not clear what a good level of EPR is, but our key findings provide new insights: (1) OFF-medication EPR level in PwPD is higher than that of HCs and (2) DRT tends to bring EPR down. This suggests that healthy brain function requires an optimal level of EPR balancing random and structured activity. Brain diseases such as Alzheimer’s disease reduce EPR^24^ while PD seem to increase EPR (this study). Clearly, further work is needed to characterize EPR changes in other brain diseases and to understand the underlying mechanisms leading to diverging effects of different forms of neurodegeneration on ERP.

This, to the best of our knowledge, is the first systematic analysis of the effect of PD and its treatment with DRT on EPR. Our results underscore the importance of studying non-equilibrium dynamics of the brain using model-based methods such as EPR to better understand the pathophysiology of neurodegenerative diseases and effects of treatment.

## Data Availability

Due to privacy concerns regarding the identities of the persons with Parkinson's disease, the data is exclusively accessible in Stockholm and only by personnel verified by the NatMEG.

## Contributors

## Declaration of interests

We declare no competing interests.

## Acknowledgments

We thank Matthieu Gilson for insightful discussions on the interpretation of EPR from a modelling perspective.

## Notes

### Competing Interest Statement

The authors have declared no competing interest.

### Funding Statement

Digital Futures, StratNeuro, NatMEG experiments were funded by Stiftelsen for Strategisk Forskning, Swedish Research Council Grant No. 638-2013-9243.

### Author Declarations

We received approval for this study from the the Regional Ethical Review Board in Stockholm (Regionala Etikprövningsnä;mnden i Stockholm, DNR 2016/911-31/1, last updated with amendment DNR 2024-00658-02) with affiliation: the Swedish Ethical Review Authority (Etikprövningsmyndigheten).

## References

1 McGregor MM, Nelson AB. Circuit Mechanisms of Parkinson’s Disease. Neuron. 2019;101(6):1042–56.

2 Geraedts VJ, Boon LI, Marinus J, Gouw AA, van Hilten JJ, Stam CJ, et al. Clinical correlates of quantitative EEG in Parkinson disease: A systematic review. Neurology. 2018;91(19):871–83.

3 Boon LI, Geraedts VJ, Hillebrand A, Tannemaat MR, Contarino MF, Stam CJ, et al. A systematic review of MEG-based studies in Parkinson’s disease: The motor system and beyond. Human Brain Mapping. 2019;40(9):2827–48.

4 Brunel N. Dynamics of Sparsely Connected Networks of Excitatory and Inhibitory Spiking Neurons. Journal of Computational Neuroscience. 2000;8(3):183–208.

5 Nartallo-Kaluarachchi R, Kringelbach ML, Deco G, Lambiotte R, Goriely A. Nonequilibrium physics of brain dynamics. arXiv preprint arXiv:250412188. 2025.

6 Schrödinger E. What is life?: With mind and matter and autobiographical sketches. Cambridge university press; 1958.

7 Seifert U. Stochastic thermodynamics, fluctuation theorems, and molecular machines. Reports on Progress in Physics. 2012;75(12):126001.

8 Manikandan SK, Gupta D, Krishnamurthy S. Inferring Entropy Production from Short Experiments. Physical Review Letters. 2020;124(12):120603.

9 Helson P, Lundqvist D, Svenningsson P, Vinding MC, Kumar A. Cortex-wide topography of 1/f-exponent in Parkinson’s disease. npj Parkinson’s Disease. 2023;9(1):1–11.

10 Vinding MC, Tsitsi P, Waldthaler J, Oostenveld R, Ingvar M, Svenningsson P, et al. Reduction of spontaneous cortical beta bursts in Parkinson’s disease is linked to symptom severity. Brain Communications. 2020;2(1):fcaa052.

11 Gramfort A, Luessi M, Larson E, Engemann DA, Strohmeier D, Brodbeck C, et al. MEG and EEG data analysis with MNE-Python. Frontiers in Neuroscience. 2013;7.

12 Glasser MF, Coalson TS, Robinson EC, Hacker CD, Harwell J, Yacoub E, et al. A multi-modal parcellation of human cerebral cortex. Nature. 2016;536(7615):171–8.

13 Jarzynski C. Equalities and inequalities: Irreversibility and the second law of thermodynamics at the nanoscale. Progress in Mathematical Physics. 2013 01;63:145–72.

14 Barato AC, Seifert U. Thermodynamic uncertainty relation for biomolecular processes. Physical review letters. 2015;114(15):158101.

15 Goetz CG, Tilley BC, Shaftman SR, Stebbins GT, Fahn S, Martinez-Martin P, et al. Movement Disorder Society-sponsored revision of the Unified Parkinson’s Disease Rating Scale (MDS-UPDRS): scale presentation and clinimetric testing results. Movement disorders: official journal of the Movement Disorder Society. 2008;23(15):2129–70.

16 Otsubo S, Ito S, Dechant A, Sagawa T. Estimating entropy production by machine learning of short-time fluctuating currents. Physical Review E. 2020;101(6):062106.

17 Airaksinen K, Butorina A, Pekkonen E, Nurminen J, Taulu S, Ahonen A, et al. Somatomotor mu rhythm amplitude correlates with rigidity during deep brain stimulation in Parkinsonian patients. Clinical Neurophysiology. 2012;123(10):2010–7.

18 Little S, Brown P. The functional role of beta oscillations in Parkinson’s disease. Parkinsonism & related disorders. 2014;20:S44–8.

19 Pauls KAM, Korsun O, Nenonen J, Nurminen J, Liljeström M, Kujala J, et al. Cortical beta burst dynamics are altered in Parkinson’s disease but normalized by deep brain stimulation. NeuroImage. 2022;257:119308.

20 Vinding MC, Waldthaler J, Eriksson A, Manting CL, Ferreira D, Ingvar M, et al. Oscillatory and nonoscillatory features of the magnetoencephalic sensorimotor rhythm in Parkinson’s disease. npj Parkinson’s Disease. 2024;10(1):51.

21 De Benedictis A, Duffau H, Paradiso B, Grandi E, Balbi S, Granieri E, et al. Anatomo-functional study of the temporo-parieto-occipital region: dissection, tractographic and brain mapping evidence from a neurosurgical perspective. Journal of anatomy. 2014;225(2):132–51.

22 Wiesman AI, Vinding MC, Tsitsi P, Svenningsson P, Waldthaler J, Lundqvist D. Cortical effects of dopamine replacement account for clinical response variability in Parkinson’s disease. Movement Disorders. 2025.

23 Surmeier DJ, Obeso JA, Halliday GM. Selective neuronal vulnerability in Parkinson disease. Nature Reviews Neuroscience. 2017;18(2):101–13.

24 Cruzat J, Herzog R, Prado P, Sanz-Perl Y, Gonzalez-Gomez R, Moguilner S, et al. Temporal Irreversibility of Large-Scale Brain Dynamics in Alzheimer’s Disease. The Journal of Neuroscience. 2023;43(9):1643–56.

25 Wiest C, Torrecillos F, Pogosyan A, Bange M, Muthuraman M, Groppa S, et al. The aperiodic exponent of subthalamic field potentials reflects excitation/inhibition balance in Parkinsonism. elife. 2023;12:e82467.

26 van Wijk B, Jha A, Penny W, Litvak V. Parametric estimation of cross-frequency coupling. Journal of neuroscience methods. 2015;243:94–102.

27 Tinkhauser G, Pogosyan A, Tan H, Herz DM, Kühn AA, Brown P. Beta burst dynamics in Parkinson’s disease OFF and ON dopaminergic medication. Brain. 2017;140(11):2968–81.

28 Byrne A, Brookes MJ, Coombes S. A mean field model for movement induced changes in the beta rhythm. Journal of computational neuroscience. 2017;43(2):143–58.

29 Manikandan SK, Ghosh T, Mandal T, Biswas A, Sinha B, Mitra D. Estimate of entropy production rate can spatiotemporally resolve the active nature of cell flickering. Physical Review Research. 2024;6(2):023310.

30 Manikandan SK, Ghosh S, Kundu A, Das B, Agrawal V, Mitra D, et al. Quantitative analysis of non-equilibrium systems from short-time experimental data. Communications Physics. 2021;4(1):1–10.

31 Das B, Manikandan SK. Localizing entropy production along non-equilibrium trajectories. arXiv preprint arXiv:250320427. 2025.

32 Van Vu T, Vo VT, Hasegawa Y. Entropy production estimation with optimal current. Physical Review E. 2020;101(4):042138.

33 Hoehn MM, Yahr MD, et al. Parkinsonism: onset, progression, and mortality. Neurology. 1998;50(2).

