## Supplementary Information for "Non-equilibrium dynamics of the neocortex in Parkinson’s disease"

The appendix contains the results from the additional tests described in Results of the main article and some more details about the methods (especially the EPR estimation algorithm).

#### EPR estimation algorithm

We compute entropy production rate by solving the optimisation problem

$$\sigma = \max_{J_{dt}} \frac{2k_B}{dt} \frac{\langle J_{dt} \rangle_t^2}{Var_t(J_{dt})}, \tag{.1}$$

where $J_{dt}(t) = \mathbf{d}(\frac{\mathbf{x}(t)+\mathbf{x}(t+dt)}{2}) \cdot (\mathbf{x}(t+dt) - \mathbf{x}(t))$ is a scalar and $\mathbf{d}(\mathbf{x})$ a vector. The maximisation is performed over the function $\mathbf{d}$, which is defined by a neural network and optimised using standard machine learning techniques as discussed in.[29,30,31] Note that we used the ergodic property of our signal to replace the statistics over randomness in (1) by the statistics over time in (2). It has been shown that the optimal bound is obtained in the short-time limit, $\tau \to 0$ in (1), for some specific models – namely overdamped Langevin systems.[8,32,16]

Our estimator for the EPR is a solution of an optimisation problem. Hence, the results might depend on the solver that we used – here a neural network as well as a linear optimiser using gradient descent (*pyswarm* package). Even though we did not see significant differences between the methods we used, a more systematic analysis would be needed. In addition, the EPR can be estimated in other ways, as referred to in the introduction. A comparison of different methods to estimate EPR is thus also needed.

### Supplementary Figures

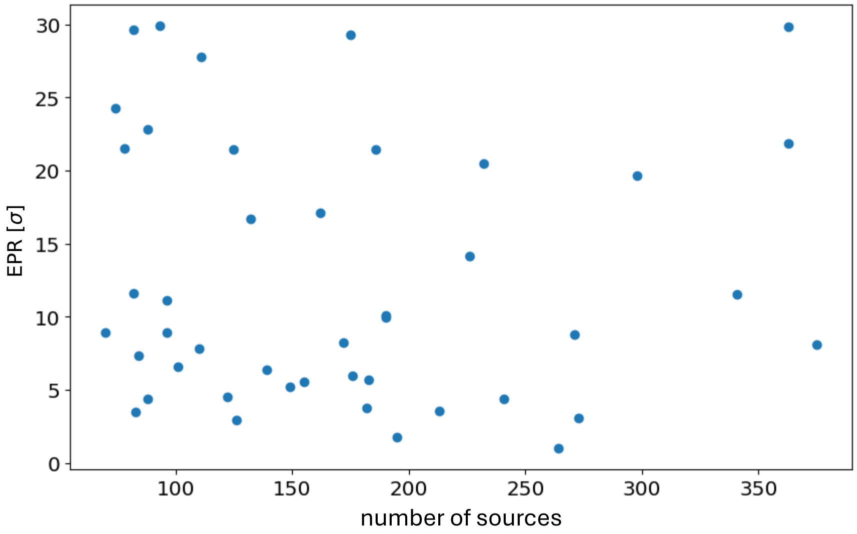


Figure S1: **EPR with respect to the number of sources.**

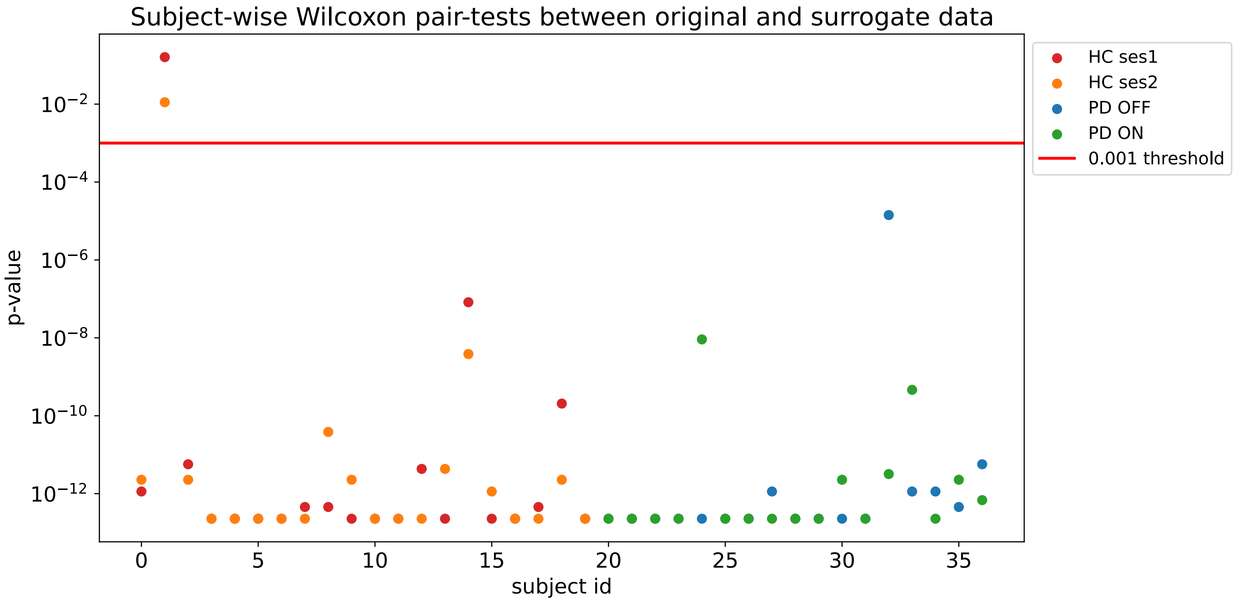


Figure S2: **Subject-wise Wilcoxon pair-tests between original and surrogate data**

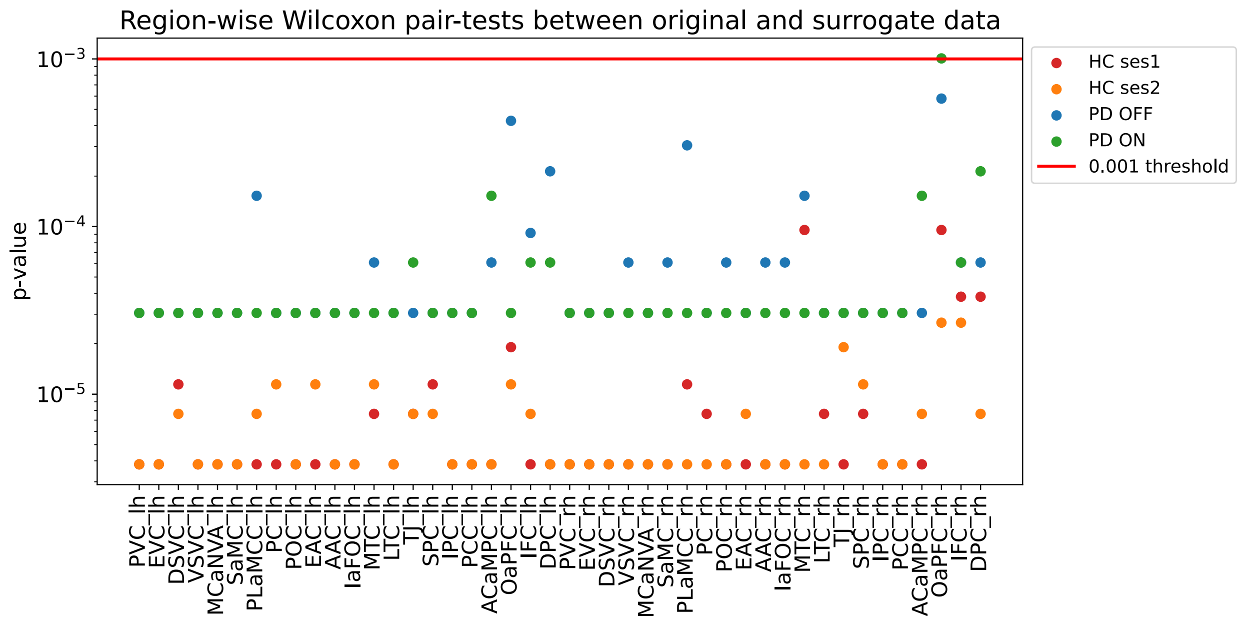


Figure S3: **Region-wise Wilcoxon pair-tests between original and surrogate data**

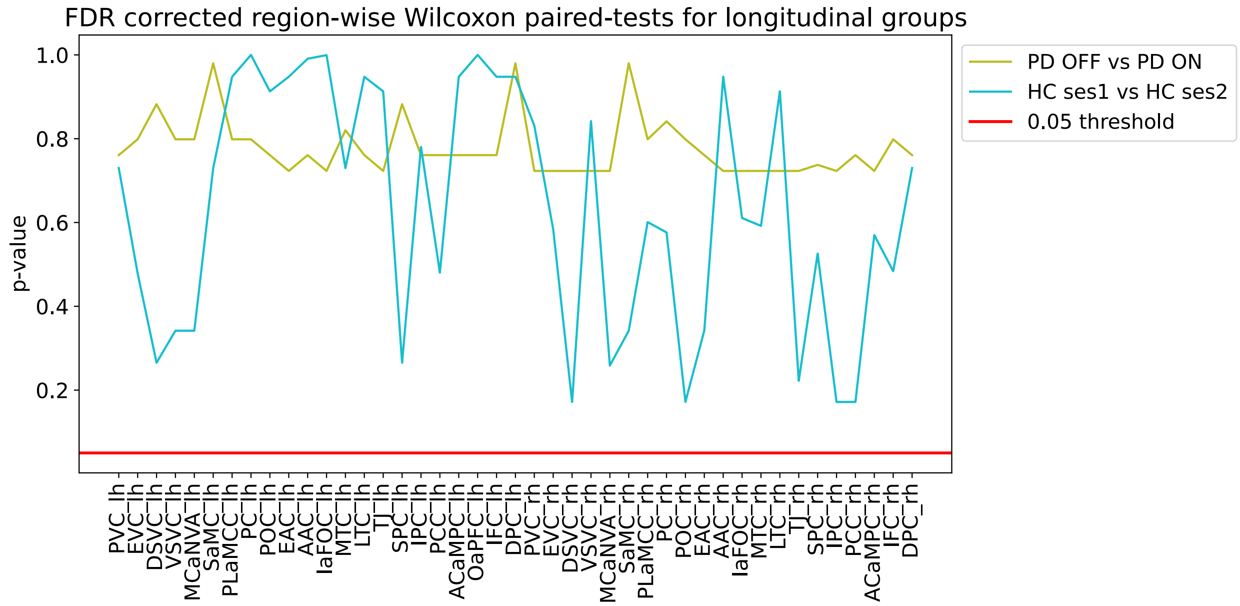


Figure S4: **FDR corrected region-wise Wilcoxon paired-tests for longitudinal groups**

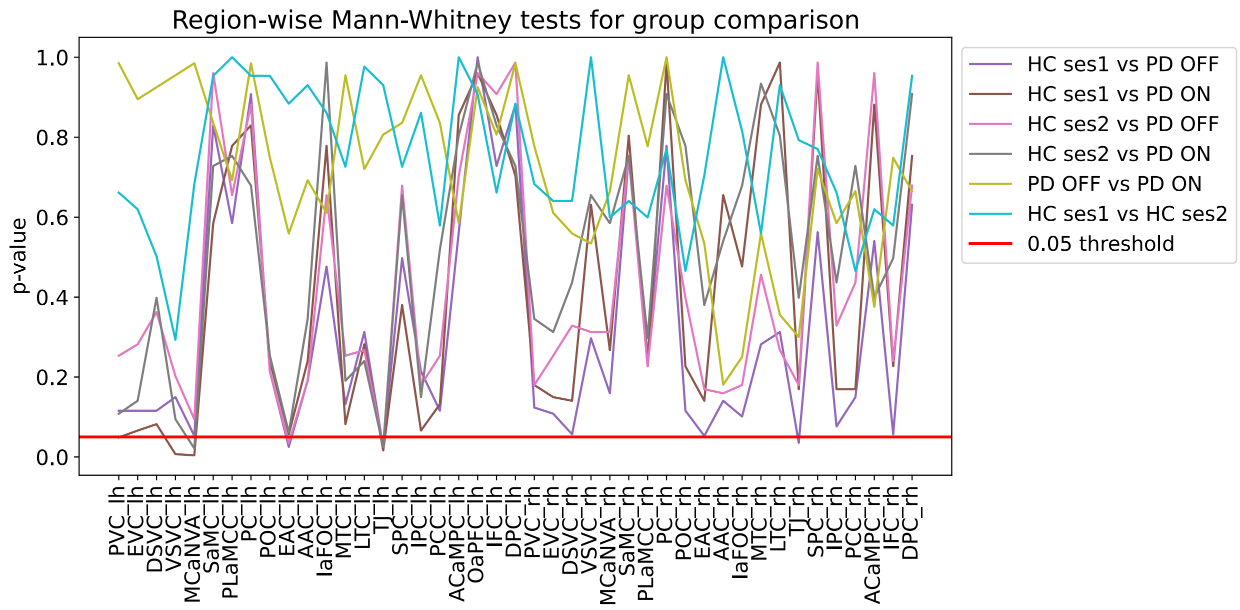


Figure S5: **Region-wise Mann-Whitney tests for group comparison before FDR correction**

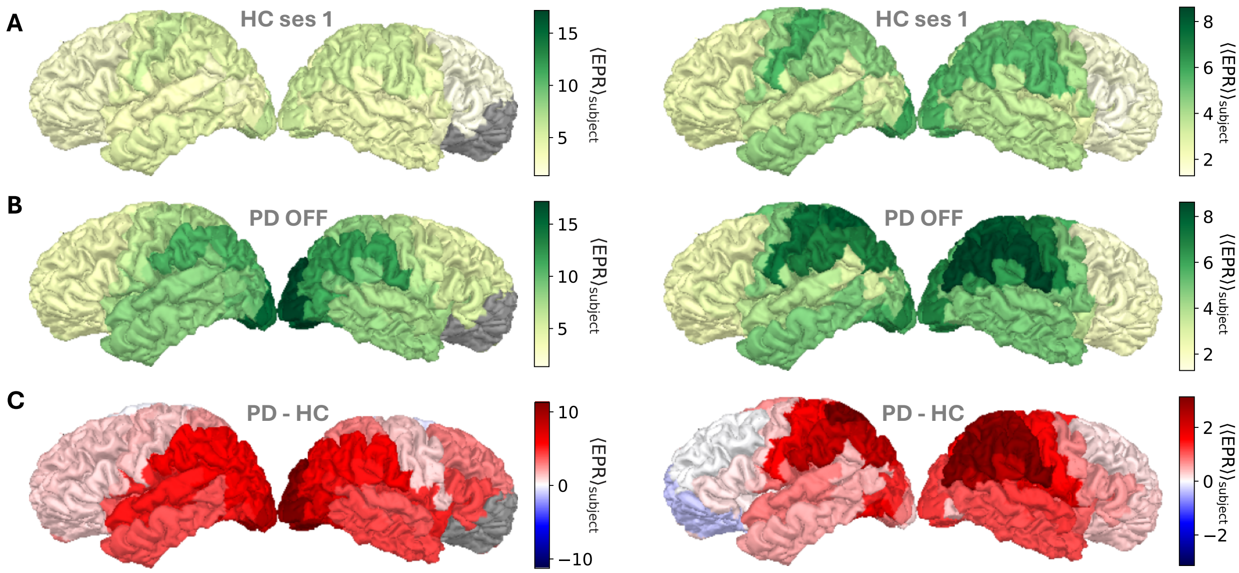


Figure S6: **Brain visualisation of the EPR statistics over the groups.** The EPR mean (left, $\langle \text{EPR} \rangle_{\text{subject}}$) and SD (right, $\langle\langle \text{EPR} \rangle\rangle_{\text{subject}}$) over different groups – **(A)** HC session 1 and **(B)** PD OFF – for each brain region. **(C)** The difference between PD OFF and HC session 1.

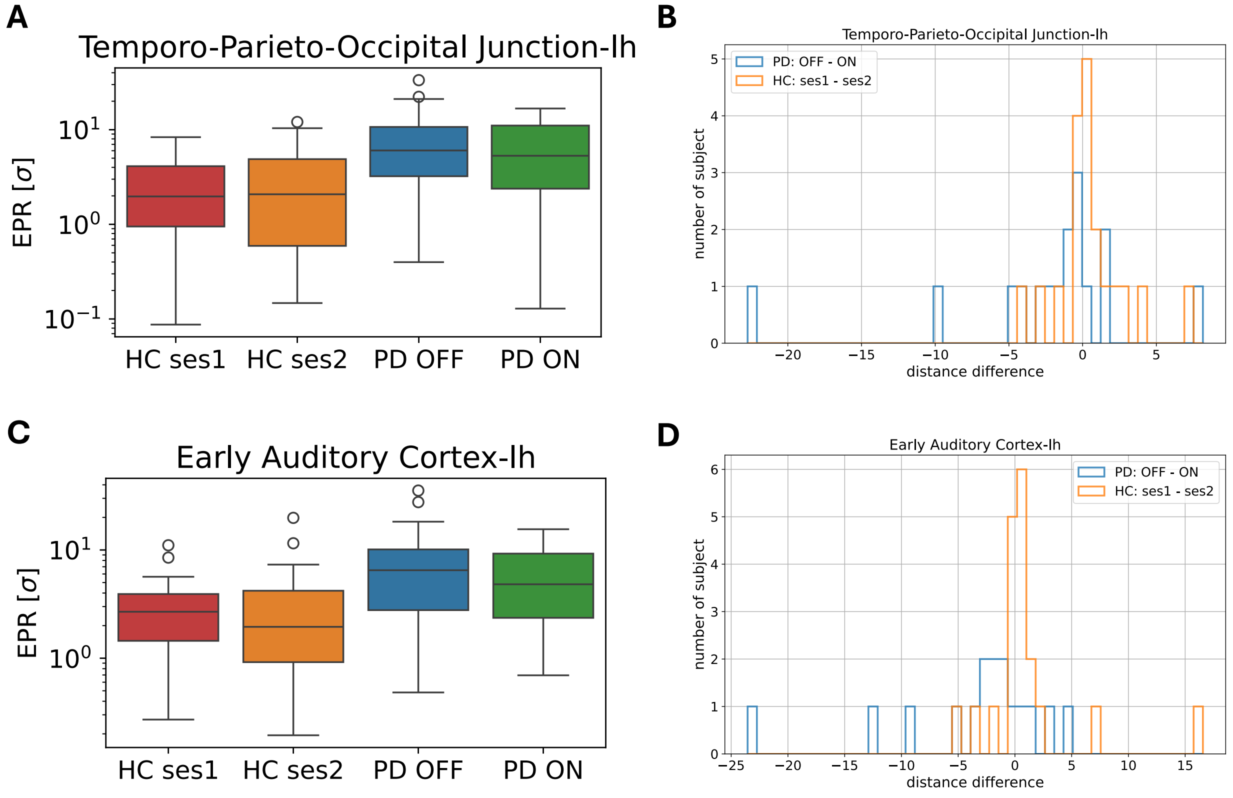


Figure S7: **Box plots of the two brain regions with the lowest p-values for the permutation** (200000 **permutations) test HC ses1 vs PD OFF.** The original p-values are 0.0037 for the temporo-parieto-occipital junction region and 0.0057 for the early auditory cortex. Both are at 0.088 after FDR correction.

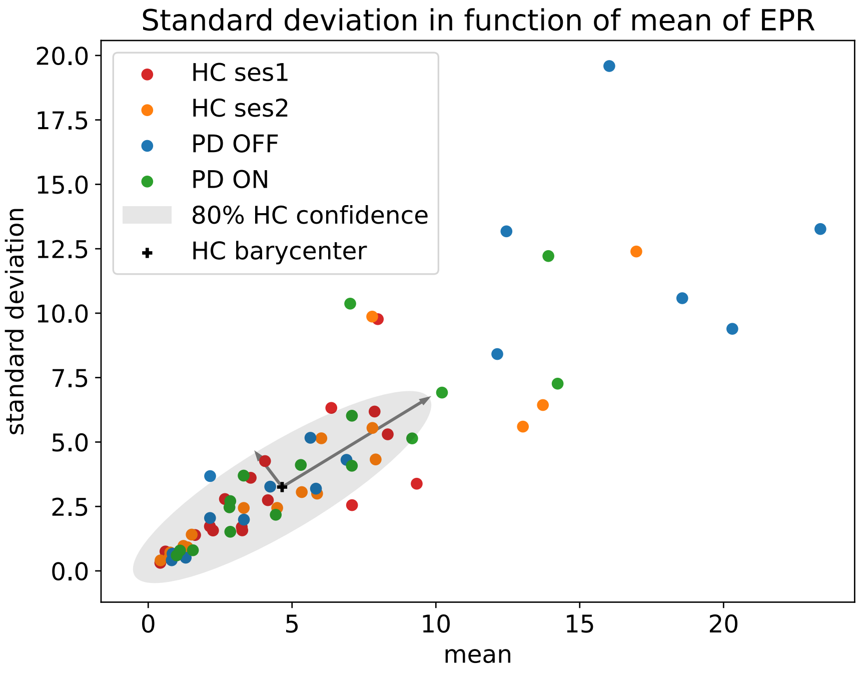


Figure S8: **Spatial standard deviation in function of mean of EPR**

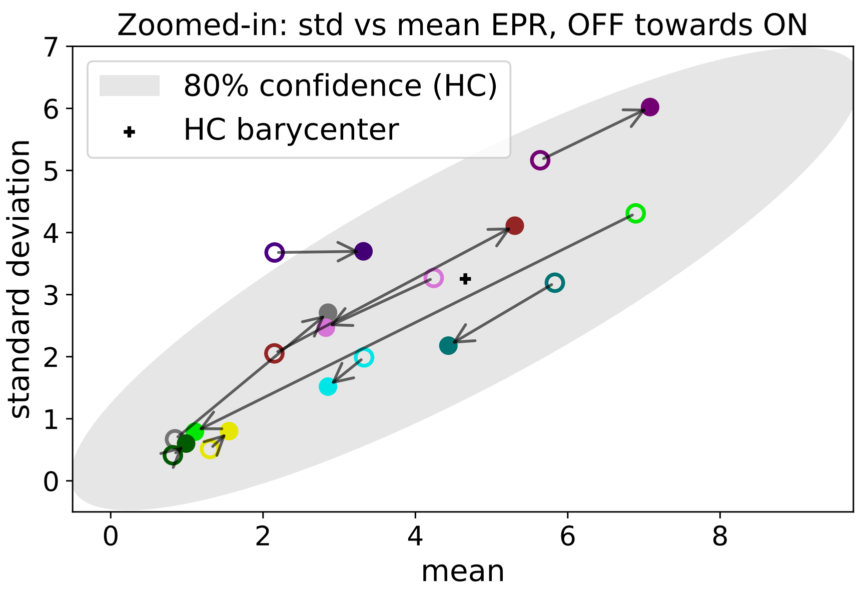


Figure S9: **Zoomed-in: std vs mean EPR, OFF towards ON**

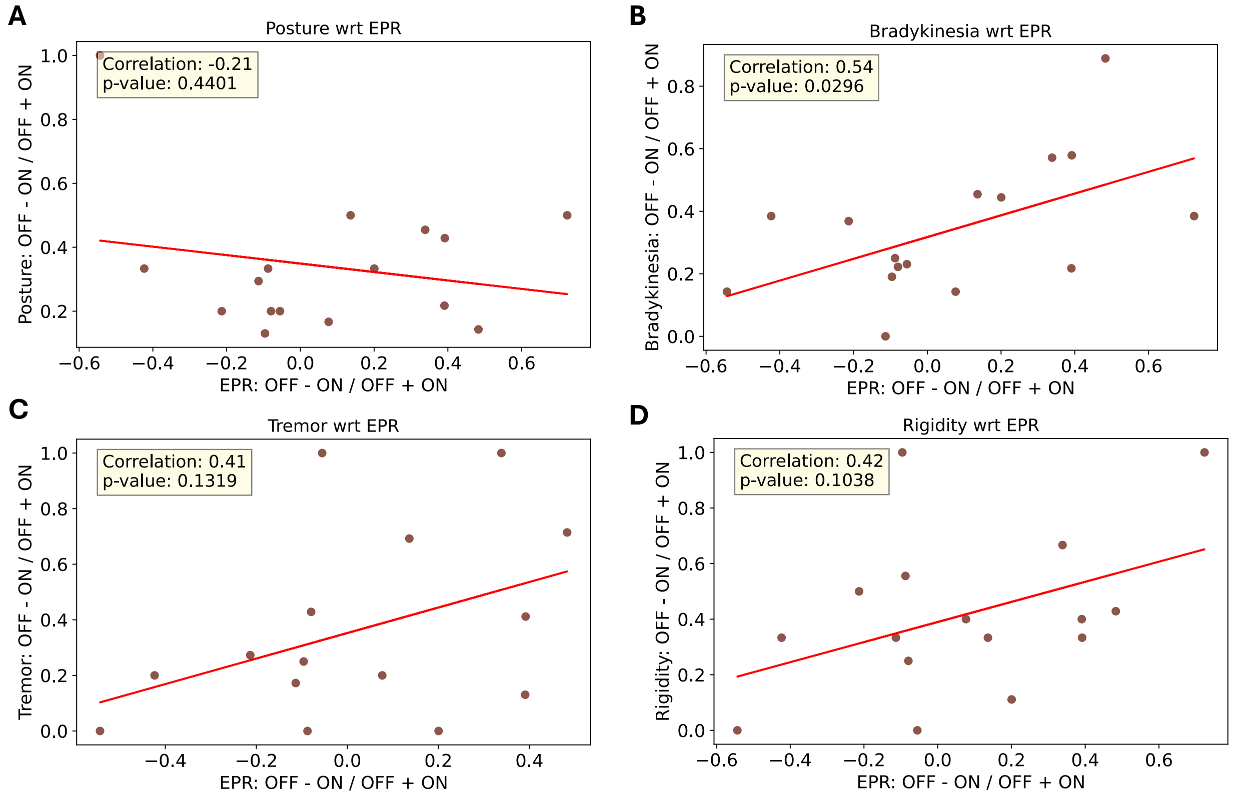


Figure S10: **Correlations between the relative motor symptoms and the relative EPR.** Gathering the UPDRS-III scores of Goetz et al.[15] as posture = {F1}, bradykinesia = {F4, F5, F7}, tremor = {F2, F6}, rigidity = {F3}.

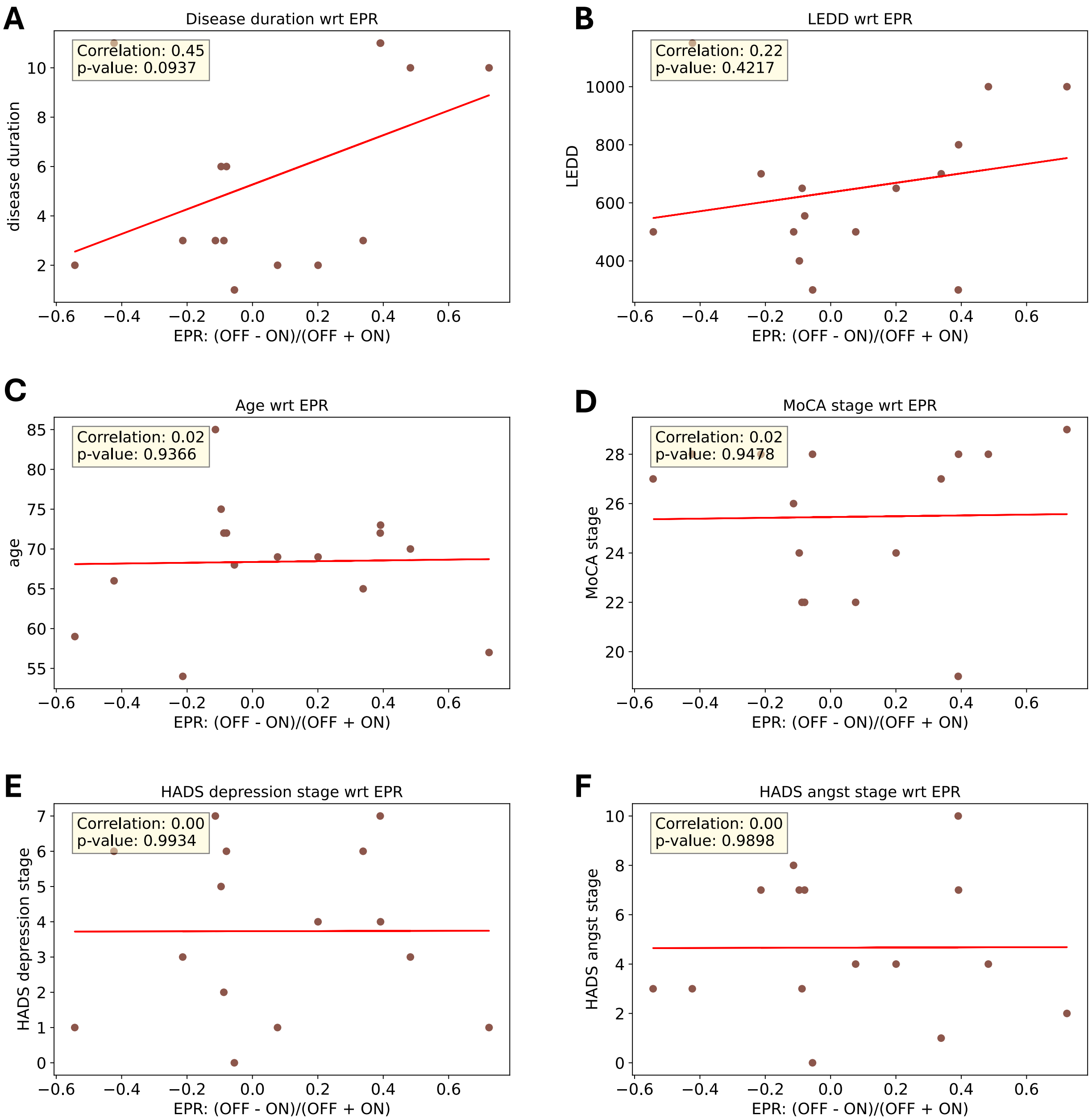


Figure S11: **Correlations between the different variables and the relative EPR.**

| VARIABLE / GROUP | Age (year) | LEDD (mg) | Disease duration (year) | HY stage | UPDRS-III | | | | | | |
|---|---|---|---|---|---|---|---|---|---|---|---|
| | | | | | F1 Gait | F2 R tremor | F3 Rigidity | F4-5 Hand | F6 P-K tremor | F7 Legs | TOTAL |
| HC | 69,4 ± 5,7 | NA | NA | 0 | 0,4 ± 0,8 | 0,1 ± 0,4 | 0,2 ± 0,5 | 0,6 ± 1 | 0,2 ± 0,4 | 0,1 ± 0,4 | 1,6 ± 2,4 |
| PD ON | 67,2 ± 9,4 | 633,7 ± 280 | 5,3 ± 3,9 | 2,3 ± 0,6 | 4,9 ± 2,9 | 1,6 ± 2,6 | 2,2 ± 1,3 | 4,9 ± 3,5 | 2,3 ± 1,9 | 1,6 ± 1,5 | 17,5 ± 9,6 |
| PD OFF | | | | | 8,7 ± 3,9 | 4,1 ± 4,3 | 4,9 ± 2,1 | 8,4 ± 3,8 | 3,4 ± 2,6 | 3,8 ± 2,8 | 33,2 ± 11,6 |

Table T1: **PwPD and HC details.** This table indicates the mean ± SD of the different participant information. Hoehn and Yahr (HY) stage is a measure of PD progress.[33] R (resp. P-K) tremor means resting (resp. postural and kinetic) tremor. For more details on MDS-UPDRS-III subscales (F1-7) we refer to Goetz et al.[15]

| | PwPD1 | PwPD2 | PwPD3 | PwPD4 | PwPD5 | PwPD6 | PwPD7 | PwPD8 | PwPD9 | PwPD10 | PwPD11 | PwPD12 | PwPD13 | PwPD14 | PwPD15 | PwPD16 |
|---|---|---|---|---|---|---|---|---|---|---|---|---|---|---|---|---|
| PVC_lh | 23 | 14 | 48 | 0.35 | 12 | 16 | 1.1 | 0.73 | 2.2 | 2.3 | 3.2 | 7.7 | 5.7 | 42 | 48 | 1.2 |
| EVC_lh | 28 | 18 | 50 | 0.73 | 15 | 17 | 0.91 | 0.87 | 2.3 | 2.3 | 4.3 | 7.8 | 7.9 | 37 | 49 | 0.99 |
| DSVC_lh | 5.5 | 7.5 | 30 | 0.36 | 14 | 12 | 0.35 | 1.3 | 3.3 | 0.83 | 3.7 | 3.5 | 5.4 | 26 | 24 | 0.88 |
| VSVC_lh | 27 | 20 | 20 | 1.8 | 9.4 | 21 | 0.51 | 0.65 | 2.5 | 2.5 | 1.5 | 2.6 | 9.9 | 12 | 14 | 0.74 |
| MCaNVA_lh | 20 | 20 | 29 | 0.96 | 11 | 20 | 0.64 | 0.95 | 3.1 | 2.3 | 2.7 | 1.9 | 9.7 | 23 | 11 | 0.93 |
| SaMC_lh | 20 | 7.4 | 31 | 2.4 | 1.4 | 17 | 1.2 | 2.5 | 5.3 | 4.1 | 11 | 0.17 | 3.4 | 8.5 | 0.47 | 0.92 |
| PLaMCC_lh | 5.7 | 11 | 18 | 2 | 0.3 | 5.2 | 0.37 | 2.1 | 2.6 | 1.9 | 12 | 0.1 | 1.7 | 8.6 | 0.049 | 0.63 |
| PC_lh | 16 | 16 | 21 | 2 | 0.5 | 9.6 | 0.26 | 3.2 | 3.4 | 4.2 | 13 | 0.31 | 1.7 | 5.8 | 0.28 | 0.9 |
| POC_lh | 29 | 14 | 14 | 1.4 | 3.8 | 37 | 4.2 | 0.72 | 4.3 | 7.2 | 6.9 | 0.4 | 3.6 | 3 | 2.4 | 0.93 |
| EAC_lh | 28 | 13 | 18 | 1.8 | 6.5 | 35 | 5.4 | 0.64 | 3.1 | 9.3 | 8.2 | 0.48 | 6.5 | 3.6 | 9.2 | 0.97 |
| AAC_lh | 22 | 11 | 19 | 1.5 | 3.6 | 36 | 1.6 | 0.77 | 2.4 | 16 | 5.3 | 0.39 | 3.7 | 3 | 9.6 | 0.94 |
| IaFOC_lh | 22 | 17 | 13 | 1.4 | 2.5 | 28 | 2 | 0.75 | 3.1 | 9.8 | 4.3 | 0.26 | 0.56 | 2 | 11 | 1.1 |
| MTC_lh | 15 | 13 | 12 | 1.8 | 3.6 | 26 | 0.99 | 0.77 | 1.6 | 5.2 | 2.3 | 0.21 | 4.1 | 6.2 | 17 | 0.73 |
| LTC_lh | 20 | 19 | 17 | 1.8 | 5.8 | 27 | 1.2 | 0.59 | 1.7 | 14 | 3.7 | 0.31 | 3.2 | 4 | 14 | 1.2 |
| TJ_lh | 21 | 13 | 22 | 1.1 | 6.9 | 33 | 4.1 | 0.4 | 3.9 | 4 | 8.6 | 0.69 | 10 | 6.5 | 5.6 | 0.6 |
| SPC_lh | 14 | 5.2 | 32 | 1.3 | 11 | 11 | 2.7 | 0.69 | 3 | 2.4 | 8.3 | 0.32 | 4.8 | 13 | 1.9 | 0.59 |
| IPC_lh | 28 | 13 | 35 | 2.1 | 13 | 35 | 5 | 0.64 | 5.3 | 4 | 9 | 0.57 | 9.6 | 10 | 2.1 | 0.65 |
| PCC_lh | 18 | 16 | 36 | 1.5 | 18 | 23 | 1.9 | 1.7 | 4.4 | 1.9 | 13 | 1.5 | 6.5 | 25 | 17 | 0.63 |
| ACaMPC_lh | 5.5 | 29 | 6.5 | 1.5 | 0.15 | 5 | 0.8 | 0.61 | 1.9 | 8.1 | 11 | 0.035 | 0.26 | 1.1 | 5.4 | 0.25 |
| OaPFC_lh | 9.3 | 21 | 3.7 | 0.86 | 0.081 | 6.1 | 0.75 | 0.34 | 0.56 | 7.7 | 1.5 | 0.025 | 0.32 | 0.12 | 7.6 | 0.25 |
| IFC_lh | 11 | 13 | 5.7 | 1.7 | 0.28 | 7.8 | 1.4 | 0.69 | 1 | 9.8 | 4.4 | 0.037 | 0.18 | 0.39 | 2.9 | 0.5 |
| DPC_lh | 2.8 | 21 | 6.3 | 2 | 0.1 | 2 | 1.4 | 0.96 | 1.3 | 8.3 | 9.3 | 0.065 | 0.036 | 0.51 | 2.8 | 0.17 |
| PVC_rh | 10 | 33 | 39 | 0.79 | 14 | 9.2 | 2.3 | 1.1 | 2.2 | 3.7 | 2.7 | 14 | 6.3 | 44 | 68 | 1.5 |
| EVC_rh | 8.5 | 35 | 45 | 0.62 | 14 | 14 | 2 | 0.78 | 3.3 | 4.7 | 3.4 | 14 | 9.6 | 45 | 73 | 1.4 |
| DSVC_rh | 4.9 | 10 | 32 | 0.75 | 7.1 | 5.9 | 1.5 | 0.72 | 6.5 | 3.1 | 1.8 | 6.6 | 8.1 | 28 | 62 | 0.92 |
| VSVC_rh | 6.1 | 38 | 24 | 0.76 | 4.9 | 16 | 1.1 | 0.45 | 0.73 | 3.9 | 6.6 | 7.7 | 6.8 | 32 | 42 | 0.11 |
| MCaNVA_rh | 3.6 | 27 | 33 | 0.92 | 7 | 13 | 0.67 | 0.23 | 1.5 | 4.4 | 4.3 | 8.9 | 7.5 | 25 | 45 | 0.53 |
| SaMC_rh | 8 | 14 | 37 | 0.97 | 2.7 | 16 | 1.5 | 0.18 | 5.6 | 11 | 6.2 | 0.086 | 1.4 | 5.1 | 1.5 | 0.94 |
| PLaMCC_rh | 3.7 | 13 | 19 | 0.75 | 0.57 | 3.5 | 0.036 | 0.16 | 5 | 3.2 | 3.8 | 0.061 | 1.6 | 5.5 | 0.47 | 1.3 |
| PC_rh | 5.2 | 20 | 21 | 1.5 | 1.3 | 13 | 2.8 | 0.27 | 4.2 | 11 | 2.1 | 0.067 | 0.45 | 1.2 | 1 | 1.6 |
| POC_rh | 8.6 | 19 | 21 | 0.67 | 3.9 | 34 | 4.4 | 0.54 | 5.9 | 13 | 5.3 | 0.021 | 1.6 | 2 | 2.9 | 1.5 |
| EAC_rh | 9.2 | 22 | 18 | 1 | 3.8 | 31 | 3 | 0.77 | 4.7 | 9.2 | 6.2 | 0.18 | 2.7 | 3.8 | 4.1 | 1.2 |
| AAC_rh | 5.4 | 29 | 9.4 | 1.5 | 2.1 | 28 | 4.4 | 2.3 | 4.1 | 12 | 5.2 | 0.12 | 2.7 | 3.7 | 7.4 | 1.6 |
| IaFOC_rh | 6.3 | 29 | 12 | 1.8 | 2.7 | 30 | 7 | 1.2 | 3.9 | 16 | 3.1 | 0.023 | 0.76 | 1.5 | 5.9 | 1.2 |
| MTC_rh | 4 | 36 | 15 | 0.79 | 2.5 | 21 | 0.26 | 0.88 | 0.79 | 5.2 | 4.1 | 1.3 | 3.2 | 11 | 14 | 0.17 |
| LTC_rh | 3.7 | 39 | 14 | 1.4 | 2.7 | 23 | 1.3 | 1.9 | 1.3 | 7.7 | 5.1 | 0.69 | 2.6 | 11 | 14 | 0.79 |
| TJ_rh | 7.8 | 21 | 32 | 0.72 | 5.2 | 31 | 2.1 | 0.43 | 4.3 | 6.2 | 6.3 | 3.3 | 4.9 | 15 | 7.8 | 0.7 |
| SPC_rh | 10 | 10 | 41 | 0.87 | 4.8 | 11 | 0.9 | 0.27 | 8.7 | 3.8 | 7.2 | 0.2 | 7.3 | 10 | 17 | 0.24 |
| IPC_rh | 12 | 17 | 45 | 1.3 | 6.9 | 28 | 2.3 | 0.38 | 7.5 | 9.1 | 8.7 | 1.6 | 6.7 | 16 | 28 | 0.86 |
| PCC_rh | 8.4 | 26 | 42 | 1.6 | 14 | 20 | 1.8 | 0.58 | 7.2 | 2.2 | 6.7 | 3.4 | 8.5 | 32 | 35 | 0.35 |
| ACaMPC_rh | 3.1 | 34 | 3.8 | 1.8 | 0.25 | 5.9 | 0.81 | 0.26 | 0.89 | 12 | 7.9 | 0.13 | 0.23 | 1.2 | 3.5 | 0.3 |
| IFC_rh | 2.6 | 32 | 5.5 | 1.4 | 0.77 | 11 | 11 | 0.27 | 1.4 | 15 | 1.7 | 0.071 | 0.27 | 0.39 | 1.9 | 0.32 |
| DPC_rh | 1 | 38 | 4.5 | 1.6 | 0.6 | 4.2 | 2.8 | 0.15 | 1.1 | 11 | 4.5 | 0.049 | 0.29 | 0.86 | 0.41 | 0.78 |

Table T2: **EPR of PD OFF across brain regions.**

| | PwPD1 | PwPD2 | PwPD3 | PwPD4 | PwPD5 | PwPD6 | PwPD7 | PwPD8 | PwPD9 | PwPD10 | PwPD11 | PwPD12 | PwPD13 | PwPD14 | PwPD15 | PwPD16 |
|---|---|---|---|---|---|---|---|---|---|---|---|---|---|---|---|---|
| PVC_lh | 26 | 8.6 | 17 | 0.22 | 16 | 8.2 | 7.9 | 9.5 | 2.2 | 1.2 | 3.9 | 12 | 4.5 | 37 | 19 | 1 |
| EVC_lh | 33 | 10 | 17 | 1.3 | 19 | 8.3 | 8.2 | 10 | 2 | 0.97 | 4.7 | 14 | 5.7 | 35 | 23 | 0.82 |
| DSVC_lh | 11 | 3.2 | 12 | 0.66 | 17 | 4.8 | 1.2 | 6.5 | 1.8 | 0.19 | 3.4 | 8.1 | 4.6 | 27 | 8 | 1.2 |
| VSVC_lh | 34 | 9.7 | 7.1 | 2.8 | 13 | 8.9 | 13 | 6.5 | 1.7 | 0.46 | 3.5 | 5.4 | 6.1 | 9 | 6.2 | 0.95 |
| MCaNVA_lh | 25 | 12 | 14 | 1.5 | 15 | 11 | 9 | 9 | 2.3 | 0.29 | 4.3 | 7.7 | 5.7 | 16 | 7.4 | 1.3 |
| SaMC_lh | 21 | 15 | 14 | 3.2 | 2.1 | 8.3 | 2.6 | 4.8 | 2.8 | 1.1 | 11 | 4.6 | 1.7 | 8 | 0.6 | 1.9 |
| PLaMCC_lh | 10 | 9.1 | 8 | 3.4 | 0.84 | 3.3 | 0.58 | 5.5 | 2.4 | 0.44 | 6.8 | 3.1 | 0.71 | 6.8 | 0.44 | 0.72 |
| PC_lh | 18 | 11 | 9.6 | 2.4 | 1.5 | 6.5 | 1.8 | 7.2 | 2.7 | 0.16 | 8 | 3.8 | 0.33 | 6.1 | 0.44 | 2.8 |
| POC_lh | 16 | 13 | 6.1 | 2.6 | 3.1 | 7.4 | 7.8 | 0.45 | 6.2 | 0.47 | 6.8 | 3 | 1.5 | 6.2 | 1.2 | 2.3 |
| EAC_lh | 16 | 13 | 9.1 | 2.6 | 4.9 | 12 | 9.7 | 0.87 | 4.8 | 0.7 | 5.8 | 2.4 | 2.7 | 6.6 | 2.2 | 2 |
| AAC_lh | 12 | 14 | 9 | 2 | 4.6 | 15 | 10 | 0.64 | 4 | 0.43 | 4.8 | 1.7 | 1 | 6.4 | 2.8 | 0.91 |
| IaFOC_lh | 15 | 9.7 | 4.9 | 1.8 | 3.3 | 11 | 8.2 | 0.9 | 2.8 | 0.52 | 3.3 | 1.1 | 0.45 | 4.9 | 1.3 | 1.5 |
| MTC_lh | 16 | 5 | 4.5 | 2.2 | 6 | 10 | 9.3 | 2.4 | 2.1 | 1.2 | 2.5 | 1.2 | 2.1 | 6.6 | 3.4 | 1.2 |
| LTC_lh | 15 | 12 | 9.3 | 2.4 | 7.3 | 13 | 14 | 1.7 | 2.5 | 0.81 | 4 | 1.8 | 1.2 | 6.4 | 3.4 | 1.1 |
| TJ_lh | 17 | 13 | 13 | 2 | 6.5 | 11 | 12 | 3.9 | 4.1 | 0.13 | 5.4 | 2.5 | 5.2 | 8.2 | 2 | 0.97 |
| SPC_lh | 12 | 8.9 | 14 | 2.1 | 11 | 7.5 | 2.9 | 3.7 | 1.2 | 0.68 | 7.7 | 3.2 | 6.3 | 14 | 0.98 | 0.82 |
| IPC_lh | 23 | 17 | 16 | 2.8 | 9.9 | 17 | 8.3 | 3.9 | 4.4 | 0.41 | 8.7 | 3.1 | 6.2 | 12 | 1.1 | 2.3 |
| PCC_lh | 23 | 8.5 | 13 | 2.4 | 17 | 11 | 4.9 | 6.5 | 2 | 0.73 | 6.7 | 8.2 | 7.2 | 30 | 3.5 | 0.96 |
| ACaMPC_lh | 6.1 | 2.2 | 2.3 | 2.1 | 1.5 | 1.9 | 1.6 | 1.5 | 1.8 | 1.1 | 3.3 | 0.35 | 0.083 | 3.3 | 0.17 | 0.24 |
| OaPFC_lh | 5.6 | 2.9 | 1.2 | 1.6 | 2.2 | 2.1 | 5.5 | 0.2 | 1.9 | 0.97 | 1.8 | 0.16 | 0.15 | 4.6 | 0.58 | 0.31 |
| IFC_lh | 7.6 | 4.1 | 2.1 | 2.1 | 1.3 | 3.2 | 2.4 | 1 | 1.6 | 0.45 | 2.7 | 0.55 | 0.059 | 4.9 | 0.46 | 0.91 |
| DPC_lh | 6.9 | 3.7 | 3.7 | 2.3 | 0.28 | 2.4 | 1.6 | 2 | 1.6 | 0.25 | 3.3 | 0.96 | 0.04 | 4.4 | 0.49 | 0.27 |
| PVC_rh | 13 | 8.8 | 15 | 0.82 | 15 | 5.1 | 3.3 | 5.1 | 0.93 | 2.3 | 1.6 | 12 | 5.1 | 43 | 40 | 1.7 |
| EVC_rh | 9.9 | 9.4 | 18 | 0.66 | 20 | 7.1 | 3.1 | 4.4 | 1.9 | 3.2 | 2.1 | 11 | 6.9 | 45 | 39 | 1.3 |
| DSVC_rh | 9.4 | 3.9 | 19 | 0.72 | 12 | 4.6 | 0.63 | 2.5 | 2.1 | 1.1 | 1.6 | 5.8 | 4.9 | 32 | 32 | 1 |
| VSVC_rh | 6.4 | 6.3 | 6.6 | 0.84 | 5.1 | 8.7 | 1.8 | 1.7 | 1 | 1.8 | 2.6 | 3.9 | 6.3 | 33 | 20 | 0.57 |
| MCaNVA_rh | 6.8 | 5.8 | 9.3 | 0.82 | 9.1 | 9.1 | 0.62 | 1.5 | 1.2 | 1.9 | 1.7 | 3.9 | 8 | 25 | 22 | 0.7 |
| SaMC_rh | 18 | 7.3 | 21 | 0.99 | 5.1 | 12 | 2.3 | 1.4 | 5.7 | 2.7 | 6 | 0.95 | 1.2 | 5.7 | 1.4 | 0.91 |
| PLaMCC_rh | 9 | 5.9 | 12 | 0.91 | 1.3 | 3.2 | 0.1 | 2 | 6 | 0.45 | 1.6 | 1.6 | 1.1 | 4.5 | 0.85 | 0.79 |
| PC_rh | 12 | 4 | 13 | 1.2 | 4.2 | 7.4 | 5.1 | 1.1 | 3.9 | 1.5 | 6.4 | 0.21 | 0.28 | 4.2 | 1.2 | 1.1 |
| POC_rh | 18 | 5.1 | 10 | 0.79 | 6.8 | 16 | 7.9 | 0.59 | 3.8 | 2.6 | 6.9 | 0.44 | 0.4 | 5 | 2.6 | 0.57 |
| EAC_rh | 15 | 3.4 | 6.4 | 0.6 | 5.2 | 17 | 4.1 | 1.1 | 3.8 | 1.6 | 6.3 | 0.62 | 1.9 | 8.8 | 3.3 | 0.86 |
| AAC_rh | 15 | 2.4 | 2 | 0.98 | 0.75 | 15 | 8.2 | 1.2 | 2.5 | 1.2 | 4.6 | 0.45 | 2.6 | 7.8 | 4.1 | 0.87 |
| IaFOC_rh | 16 | 4 | 3.9 | 1.2 | 2.1 | 12 | 11 | 0.75 | 2.7 | 1.3 | 5.5 | 0.13 | 0.42 | 5.9 | 1.1 | 0.57 |
| MTC_rh | 4.2 | 4.2 | 3.6 | 0.76 | 1.7 | 8.3 | 1 | 1.3 | 1.9 | 1.2 | 2.2 | 0.92 | 1.9 | 16 | 4.1 | 0.39 |
| LTC_rh | 6.6 | 3.4 | 1.6 | 1.1 | 1.8 | 12 | 4.2 | 1.9 | 1.6 | 1.6 | 3.4 | 0.86 | 2.1 | 11 | 5.1 | 0.69 |
| TJ_rh | 15 | 3.2 | 9.1 | 1.1 | 7.4 | 19 | 2.7 | 0.31 | 3.5 | 2 | 3.8 | 1.5 | 3.9 | 11 | 4.4 | 0.91 |
| SPC_rh | 14 | 2.8 | 32 | 1 | 9.7 | 9.1 | 0.51 | 1.2 | 5.6 | 1.3 | 4.3 | 0.89 | 2.6 | 14 | 6.6 | 0.54 |
| IPC_rh | 22 | 3 | 26 | 1.2 | 13 | 24 | 2.4 | 0.8 | 5 | 3 | 6.3 | 1.7 | 3.1 | 18 | 12 | 1.3 |
| PCC_rh | 15 | 8.9 | 17 | 1.1 | 19 | 14 | 2.4 | 3.1 | 6 | 0.94 | 3.4 | 5.9 | 4.7 | 37 | 12 | 0.41 |
| ACaMPC_rh | 6.4 | 2.8 | 1.7 | 1.4 | 1.3 | 1.3 | 2.1 | 0.76 | 1.1 | 1.2 | 2 | 0.073 | 0.23 | 3.1 | 0.1 | 0.17 |
| IFC_rh | 6.1 | 2 | 3 | 0.86 | 1.1 | 5 | 15 | 0.42 | 1.5 | 0.66 | 3.7 | 0.3 | 0.12 | 3 | 0.41 | 0.052 |
| DPC_rh | 2.8 | 3.2 | 3.2 | 1.2 | 0.84 | 1.5 | 7.4 | 0.46 | 1.9 | 0.3 | 2.3 | 0.2 | 0.0079 | 2.7 | 0.21 | 0.53 |

Table T3: **EPR of PD ON across brain regions.**

| | PwPD1 | PwPD2 | PwPD3 | PwPD4 | PwPD5 | PwPD6 | PwPD7 | PwPD8 | PwPD9 | PwPD10 | PwPD11 | PwPD12 | PwPD13 | PwPD14 | PwPD15 | PwPD16 | PwPD17 | PwPD18 | PwPD19 |
|---|---|---|---|---|---|---|---|---|---|---|---|---|---|---|---|---|---|---|---|
| PVC_lh | 2 | 0.18 | 2.1 | 38 | 7.1 | 1.7 | 5.2 | 0.61 | 3.1 | 9.9 | 0.55 | 1.7 | 2.7 | 0.78 | 8 | 2.9 | 4.9 | 1.7 | 12 |
| EVC_lh | 2.2 | 0.45 | 3.2 | 38 | 12 | 2.2 | 5.3 | 0.85 | 4.3 | 12 | 0.72 | 1.9 | 2.2 | 0.44 | 9.6 | 3.6 | 6.5 | 1.5 | 11 |
| DSVC_lh | 0.61 | 0.19 | 1 | 4.1 | 12 | 2.6 | 1.7 | 0.67 | 1.4 | 6.9 | 0.44 | 2.8 | 1.2 | 0.0074 | 11 | 4.6 | 5.8 | 0.35 | 10 |
| VSVC_lh | 1.2 | 0.28 | 1.8 | 32 | 5 | 1.6 | 3.7 | 0.8 | 5.6 | 4.1 | 1.6 | 0.42 | 2.5 | 1.6 | 2.7 | 2.7 | 0.99 | 1.6 | 7 |
| MCaNVA_lh | 0.73 | 0.14 | 1 | 24 | 7 | 3.8 | 1.7 | 0.58 | 2.1 | 6.3 | 0.9 | 0.77 | 1.3 | 1.2 | 5.1 | 2.3 | 5 | 1.8 | 2 |
| SaMC_lh | 13 | 0.36 | 7.8 | 2 | 13 | 2.9 | 1.7 | 4.5 | 1 | 6.7 | 0.85 | 5.2 | 0.24 | 0.54 | 16 | 9.9 | 18 | 1.2 | 9.6 |
| PLaMCC_lh | 10 | 0.41 | 5.4 | 2.9 | 13 | 1.9 | 1.9 | 3.1 | 1.1 | 7.2 | 0.9 | 4.9 | 0.5 | 0.11 | 15 | 7.5 | 9.4 | 0.53 | 5.2 |
| PC_lh | 12 | 0.22 | 3 | 2.2 | 9.9 | 5.6 | 0.57 | 3.6 | 1.9 | 6.3 | 0.79 | 3.6 | 0.76 | 0.56 | 18 | 8.2 | 6.8 | 1 | 3.1 |
| POC_lh | 12 | 0.19 | 4.7 | 0.65 | 4.2 | 3.8 | 0.6 | 1 | 2.7 | 3.7 | 1.3 | 2.5 | 0.98 | 1.5 | 3.4 | 4.7 | 7.4 | 1.3 | 9.3 |
| EAC_lh | 8.5 | 0.39 | 5.7 | 1.9 | 4.7 | 2.3 | 0.27 | 0.39 | 2.7 | 3.8 | 1.9 | 1.5 | 0.52 | 2.7 | 3.2 | 4 | 3.2 | 1.4 | 11 |
| AAC_lh | 3.5 | 0.18 | 3.8 | 8.7 | 4.2 | 2.7 | 0.41 | 0.99 | 9.8 | 5 | 2 | 2.1 | 2.2 | 2.5 | 1.5 | 3.6 | 1.9 | 0.7 | 8.3 |
| IaFOC_lh | 7.9 | 0.21 | 4.3 | 3.2 | 6.4 | 6.7 | 0.5 | 3.5 | 14 | 7.6 | 0.88 | 1.3 | 2 | 1.3 | 1.3 | 3.1 | 1.9 | 0.77 | 5.5 |
| MTC_lh | 0.66 | 0.51 | 2.2 | 13 | 3.6 | 1.5 | 1.2 | 0.26 | 17 | 5.5 | 1.8 | 0.4 | 5.2 | 1.9 | 0.67 | 2.1 | 1.4 | 0.48 | 7 |
| LTC_lh | 1.2 | 0.26 | 2.3 | 26 | 7.6 | 2.9 | 1.5 | 0.81 | 15 | 5.8 | 2 | 1.9 | 4.8 | 2.9 | 1.3 | 2.7 | 1.5 | 0.69 | 5.3 |
| TJ_lh | 2.6 | 0.087 | 2.7 | 7.3 | 7.5 | 2 | 0.46 | 0.2 | 1 | 4.6 | 1.1 | 0.9 | 0.61 | 1.4 | 2.2 | 3.6 | 5.1 | 1.2 | 8.3 |
| SPC_lh | 5.1 | 0.9 | 2.9 | 1.3 | 13 | 3.1 | 1.5 | 1.6 | 0.25 | 8.8 | 0.77 | 3.5 | 0.8 | 0.025 | 12 | 9.5 | 18 | 1.4 | 9.3 |
| IPC_lh | 6 | 0.5 | 2.7 | 4.3 | 14 | 5.2 | 0.65 | 0.81 | 0.83 | 7 | 1.6 | 2.4 | 0.89 | 0.82 | 5.4 | 8.5 | 16 | 1.6 | 13 |
| PCC_lh | 5.9 | 0.66 | 4.3 | 11 | 15 | 2.2 | 2.6 | 0.79 | 1.9 | 6 | 0.85 | 3.8 | 1.5 | 0.28 | 15 | 7.8 | 8.3 | 0.92 | 20 |
| ACaMPC_lh | 1.7 | 0.06 | 1.1 | 3.6 | 6.5 | 2.9 | 0.37 | 2 | 5.9 | 6.4 | 0.94 | 1 | 4.9 | 0.099 | 3.9 | 1.8 | 0.63 | 0.14 | 1 |
| OaPFC_lh | 0.13 | 0.19 | 0.66 | 5.6 | 2.2 | 7.1 | 0.42 | 6.3 | 14 | 4.4 | 0.93 | 0.87 | 5.6 | 0.23 | 0.089 | 0.8 | 0.3 | 0.39 | 1.9 |
| IFC_lh | 6.3 | 0.22 | 1.4 | 2.1 | 5.1 | 7.9 | 0.3 | 6.6 | 7.6 | 6 | 0.51 | 0.34 | 2.4 | 0.11 | 0.32 | 2 | 0.63 | 0.33 | 1.9 |
| DPC_lh | 5 | 0.31 | 1.3 | 2.6 | 6.2 | 6.1 | 0.44 | 2.8 | 2.7 | 7.8 | 0.67 | 0.5 | 3.3 | 0.082 | 3 | 3.5 | 0.37 | 0.22 | 1.3 |
| PVC_rh | 2.1 | 0.31 | 4.5 | 12 | 9.7 | 1.6 | 4.7 | 1.2 | 2.5 | 7.4 | 0.99 | 2.9 | 3.2 | 0.57 | 17 | 3.3 | 9.4 | 1 | 14 |
| EVC_rh | 1 | 0.9 | 4.7 | 20 | 14 | 2 | 4.8 | 0.76 | 2.8 | 7.9 | 1.3 | 2.3 | 2.4 | 0.28 | 18 | 3.1 | 9.9 | 1 | 15 |
| DSVC_rh | 0.3 | 0.79 | 3.3 | 2.5 | 10 | 3.1 | 1 | 0.64 | 1.4 | 4.8 | 0.81 | 1.5 | 1 | 0.25 | 20 | 3.7 | 9.7 | 0.25 | 13 |
| VSVC_rh | 1.2 | 1 | 4.7 | 14 | 10 | 1.3 | 3.3 | 0.71 | 5.4 | 6.6 | 2.7 | 1.7 | 3.5 | 0.16 | 8 | 1.8 | 8.3 | 0.67 | 11 |
| MCaNVA_rh | 0.83 | 1 | 5.1 | 5.4 | 9.4 | 2.8 | 1.4 | 0.31 | 1.4 | 6 | 1.3 | 1.8 | 1.8 | 0.23 | 18 | 1 | 9.8 | 0.34 | 8.3 |
| SaMC_rh | 7.4 | 0.25 | 3.2 | 4.2 | 11 | 4.6 | 2.6 | 3.7 | 3.1 | 12 | 3 | 12 | 1.1 | 0.41 | 4.2 | 9.9 | 17 | 0.47 | 9.4 |
| PLaMCC_rh | 6.9 | 0.23 | 4.2 | 4.4 | 9.2 | 2.1 | 2.9 | 2.3 | 2.2 | 12 | 3.2 | 9 | 0.31 | 0.079 | 5.3 | 7 | 6.9 | 0.16 | 8.6 |
| PC_rh | 5.4 | 0.3 | 2.4 | 2.7 | 11 | 3.7 | 1.4 | 4.2 | 2.3 | 11 | 2.9 | 12 | 0.63 | 0.14 | 1.9 | 6.9 | 11 | 0.14 | 3.6 |
| POC_rh | 4.1 | 0.099 | 2.5 | 0.79 | 12 | 5.4 | 1.1 | 3.4 | 1 | 4.4 | 0.81 | 1.8 | 1.3 | 0.18 | 0.96 | 3 | 14 | 0.18 | 3.6 |
| EAC_rh | 2.4 | 0.51 | 4.3 | 1.2 | 8.1 | 2.4 | 1.6 | 2.6 | 0.84 | 6 | 1.2 | 0.67 | 2.2 | 0.29 | 0.51 | 2.4 | 14 | 0.34 | 8.7 |
| AAC_rh | 0.32 | 0.53 | 5.5 | 3.4 | 12 | 3.7 | 1.3 | 5.1 | 2.1 | 7.7 | 1.1 | 0.76 | 4 | 0.27 | 0.51 | 2.6 | 12 | 0.41 | 8.9 |
| IaFOC_rh | 0.95 | 0.075 | 2.2 | 1.9 | 11 | 4.2 | 0.73 | 5.2 | 4.2 | 4.4 | 0.77 | 0.7 | 4.2 | 0.13 | 0.87 | 1.7 | 5.7 | 0.22 | 3.1 |
| MTC_rh | 0.45 | 1.3 | 3.8 | 7.9 | 8.6 | 0.41 | 1.8 | 2.1 | 11 | 9.7 | 1.2 | 0.084 | 4.6 | 0.25 | 0.68 | 1.3 | 4.6 | 0.027 | 7.1 |
| LTC_rh | 0.41 | 0.73 | 4.6 | 9.5 | 9.4 | 1.6 | 2.1 | 4.2 | 5.5 | 10 | 2 | 1.8 | 4.6 | 0.41 | 0.98 | 1.4 | 8.8 | 0.062 | 8.5 |
| TJ_rh | 0.52 | 0.69 | 4.8 | 1.8 | 13 | 2.2 | 1.3 | 0.47 | 0.67 | 8.5 | 1 | 1.8 | 1.9 | 0.3 | 3.6 | 4.2 | 16 | 0.15 | 13 |
| SPC_rh | 2 | 0.87 | 3.3 | 3.9 | 10 | 4.5 | 2.1 | 1.6 | 1.6 | 11 | 3.5 | 7.2 | 1.4 | 0.069 | 7.8 | 8.7 | 17 | 0.18 | 17 |
| IPC_rh | 1.3 | 0.6 | 3.6 | 2.1 | 13 | 4.9 | 0.68 | 1.8 | 0.62 | 12 | 2.6 | 4.1 | 1.6 | 0.28 | 11 | 6.1 | 26 | 0.17 | 18 |
| PCC_rh | 5.9 | 0.68 | 3.8 | 4.5 | 15 | 2.3 | 1.6 | 0.7 | 2.2 | 9.7 | 2 | 5 | 0.92 | 0.26 | 14 | 7.5 | 11 | 0.65 | 22 |
| ACaMPC_rh | 0.65 | 0.058 | 1.3 | 3.7 | 9.7 | 1.9 | 0.38 | 2.3 | 5.1 | 5.6 | 1 | 1.7 | 5.6 | 0.036 | 0.85 | 1.4 | 0.23 | 0.15 | 0.55 |
| IFC_rh | 0.43 | 0.14 | 1.3 | 0.87 | 7.6 | 4 | 0.21 | 3 | 1.5 | 1.7 | 0.2 | 0.26 | 1.6 | 0.044 | 0.17 | 0.26 | 1 | 0.13 | 1.2 |
| DPC_rh | 0.45 | 0.05 | 2 | 2.4 | 6.4 | 2.3 | 0.078 | 2.9 | 1.4 | 5.1 | 0.48 | 1.4 | 2 | 0.0032 | 0.42 | 2.1 | 1.5 | 0.2 | 0.67 |

Table T4: **EPR of HC session 1 across brain regions.**

| | PwPD1 | PwPD2 | PwPD3 | PwPD4 | PwPD5 | PwPD6 | PwPD7 | PwPD8 | PwPD9 | PwPD10 | PwPD11 | PwPD12 | PwPD13 | PwPD14 | PwPD15 | PwPD16 | PwPD17 | PwPD18 | PwPD19 |
|---|---|---|---|---|---|---|---|---|---|---|---|---|---|---|---|---|---|---|---|
| PVC_lh | 2.9 | 1.4 | 2.3 | 26 | 15 | 4.5 | 6.2 | 1 | 0.71 | 9 | 0.48 | 6.3 | 2.8 | 0.29 | 7.1 | 6.3 | 8.8 | 0.5 | 9.7 |
| EVC_lh | 2.4 | 1.6 | 3.2 | 24 | 21 | 6.1 | 8 | 0.76 | 0.95 | 14 | 0.62 | 5.7 | 3 | 0.36 | 9.9 | 10 | 11 | 0.76 | 10 |
| DSVC_lh | 0.71 | 0.86 | 2.5 | 1.8 | 25 | 5.8 | 1.6 | 0.32 | 0.99 | 8.4 | 0.47 | 8.9 | 2.2 | 0.013 | 11 | 13 | 12 | 0.37 | 8.8 |
| VSVC_lh | 0.58 | 1.8 | 4.9 | 49 | 7.5 | 4.9 | 7.1 | 0.33 | 0.73 | 7.5 | 1.7 | 4.2 | 2.2 | 1.1 | 1.9 | 6.5 | 4.4 | 0.56 | 7.1 |
| MCaNVA_lh | 0.62 | 0.83 | 3 | 33 | 14 | 6.4 | 2.8 | 0.46 | 0.86 | 5.7 | 0.68 | 2.4 | 2.3 | 0.47 | 3.4 | 6.1 | 6.9 | 0.68 | 2.3 |
| SaMC_lh | 5.1 | 0.72 | 10 | 1.5 | 17 | 3.8 | 1.2 | 1.7 | 0.24 | 9.8 | 1 | 6.4 | 0.29 | 0.19 | 13 | 22 | 40 | 3.8 | 9 |
| PLaMCC_lh | 3.5 | 0.38 | 6.2 | 2.8 | 18 | 3.8 | 1.6 | 1.4 | 0.72 | 6.9 | 1.2 | 5.8 | 0.18 | 0.086 | 11 | 15 | 21 | 0.61 | 5 |
| PC_lh | 5.7 | 0.66 | 4 | 2.2 | 16 | 3.7 | 1.6 | 2.5 | 0.57 | 5.4 | 0.7 | 5.8 | 0.044 | 0.12 | 11 | 14 | 16 | 2.1 | 2.7 |
| POC_lh | 5.3 | 0.43 | 6.4 | 0.74 | 6.6 | 3.2 | 1.9 | 0.37 | 1.2 | 2.4 | 0.61 | 6 | 0.3 | 1.1 | 4.2 | 12 | 22 | 1.6 | 6.8 |
| EAC_lh | 2.7 | 0.4 | 7.3 | 2.4 | 4.8 | 1.9 | 0.83 | 0.19 | 0.58 | 1.6 | 1.1 | 3.2 | 0.62 | 1.5 | 3.6 | 12 | 20 | 1 | 7.3 |
| AAC_lh | 1.3 | 0.16 | 5.5 | 8.5 | 5.8 | 2.2 | 3.7 | 0.14 | 2.4 | 3 | 1.1 | 3.2 | 0.8 | 1.4 | 2.8 | 20 | 14 | 0.23 | 3.3 |
| IaFOC_lh | 4 | 0.78 | 5.9 | 3.8 | 5.7 | 5.7 | 7.4 | 0.41 | 1.7 | 4.1 | 0.8 | 3.7 | 0.78 | 1.3 | 4.1 | 14 | 6.7 | 0.97 | 2.7 |
| MTC_lh | 0.49 | 0.92 | 4.8 | 16 | 4.5 | 1.8 | 8.8 | 0.027 | 1.7 | 3.3 | 0.79 | 1.5 | 2.7 | 0.93 | 1.4 | 9.6 | 4.5 | 0.44 | 6.5 |
| LTC_lh | 0.94 | 0.81 | 5.1 | 26 | 11 | 2.4 | 8 | 0.14 | 2.3 | 5.6 | 0.88 | 3 | 1.4 | 1 | 2.4 | 19 | 4.9 | 0.48 | 2.4 |
| TJ_lh | 1 | 0.15 | 4.8 | 3.2 | 10 | 2.1 | 0.87 | 0.2 | 0.28 | 5 | 0.48 | 3.2 | 0.64 | 0.9 | 2.5 | 7.5 | 12 | 0.55 | 5.6 |
| SPC_lh | 1.4 | 1.4 | 6.7 | 0.93 | 25 | 6.6 | 1 | 0.82 | 0.2 | 11 | 1.5 | 8.7 | 0.93 | 0.02 | 9.4 | 24 | 44 | 2.4 | 11 |
| IPC_lh | 1.9 | 0.68 | 6.3 | 1.5 | 23 | 5.8 | 0.53 | 0.22 | 0.45 | 7.9 | 0.67 | 7.1 | 1.2 | 0.56 | 4 | 22 | 40 | 2.4 | 11 |
| PCC_lh | 2.8 | 1.6 | 7.5 | 12 | 25 | 5.5 | 1.9 | 0.63 | 0.96 | 8.5 | 1.4 | 11 | 1.2 | 0.48 | 13 | 18 | 25 | 0.6 | 17 |
| ACaMPC_lh | 0.91 | 0.81 | 1.9 | 9.9 | 7.9 | 2.6 | 5.1 | 0.88 | 0.37 | 3.7 | 0.81 | 3.1 | 0.66 | 0.061 | 3 | 5.9 | 1.4 | 0.24 | 0.5 |
| OaPFC_lh | 0.19 | 0.79 | 1.3 | 8.9 | 2.1 | 5.8 | 8.6 | 0.19 | 0.57 | 3.2 | 0.59 | 2.6 | 0.74 | 0.26 | 0.69 | 7.1 | 0.43 | 0.31 | 0.79 |
| IFC_lh | 2.2 | 0.15 | 2.9 | 1.5 | 7.2 | 8.1 | 6.2 | 0.57 | 0.95 | 3.4 | 0.86 | 3.2 | 0.45 | 0.17 | 0.98 | 12 | 3.8 | 0.37 | 1.2 |
| DPC_lh | 1.5 | 0.37 | 2.7 | 2.5 | 7.7 | 5.1 | 5.4 | 0.99 | 0.35 | 5 | 0.86 | 2.3 | 0.36 | 0.14 | 2.2 | 7.9 | 0.74 | 0.72 | 0.63 |
| PVC_rh | 1.4 | 2 | 3.8 | 7.2 | 19 | 4.2 | 4.2 | 1.6 | 0.56 | 14 | 1 | 5.3 | 1.8 | 0.42 | 14 | 8 | 21 | 0.6 | 14 |
| EVC_rh | 1.6 | 2.1 | 4.4 | 8.4 | 25 | 4.9 | 3.8 | 1.3 | 0.89 | 15 | 1.1 | 4.9 | 1.6 | 0.26 | 17 | 7.4 | 20 | 0.7 | 16 |
| DSVC_rh | 0.77 | 0.9 | 4.8 | 3.6 | 14 | 7 | 0.96 | 0.75 | 0.96 | 7.7 | 1.1 | 7.5 | 0.56 | 0.033 | 19 | 7.8 | 21 | 0.23 | 14 |
| VSVC_rh | 0.81 | 2.5 | 9.4 | 4 | 12 | 2 | 3.5 | 0.41 | 0.29 | 9.5 | 1.6 | 3.1 | 3 | 0.49 | 4.5 | 7.7 | 17 | 0.86 | 10 |
| MCaNVA_rh | 0.83 | 1.5 | 7.5 | 5.3 | 13 | 5.4 | 1.6 | 0.28 | 0.067 | 11 | 0.94 | 2.7 | 1.9 | 0.34 | 10 | 6.4 | 17 | 0.66 | 9.7 |
| SaMC_rh | 1.3 | 0.73 | 5.1 | 6.7 | 14 | 13 | 3.7 | 2.1 | 1.5 | 12 | 4.8 | 5.6 | 1.3 | 0.14 | 5.9 | 19 | 33 | 0.81 | 9.6 |
| PLaMCC_rh | 1.2 | 0.45 | 5.8 | 8 | 11 | 8.8 | 3.3 | 1 | 0.96 | 13 | 3.1 | 8 | 0.29 | 0.076 | 5.3 | 16 | 21 | 0.52 | 7.3 |
| PC_rh | 1.1 | 0.56 | 2.9 | 5.7 | 12 | 13 | 1.7 | 1.7 | 1.3 | 9.6 | 3.3 | 4.3 | 0.68 | 0.16 | 3.3 | 13 | 18 | 0.81 | 3.9 |
| POC_rh | 1.5 | 0.2 | 9 | 0.78 | 19 | 9.3 | 1.1 | 1.2 | 0.28 | 5.8 | 0.79 | 3 | 2.2 | 0.59 | 3 | 13 | 20 | 1 | 5.6 |
| EAC_rh | 0.63 | 0.4 | 11 | 1.1 | 12 | 3 | 1.6 | 0.67 | 0.046 | 7.4 | 0.65 | 2.9 | 2 | 0.75 | 2.8 | 16 | 20 | 0.47 | 10 |
| AAC_rh | 0.18 | 0.4 | 12 | 1 | 13 | 3.9 | 2.2 | 0.71 | 0.6 | 5.7 | 0.24 | 3.5 | 2.2 | 0.48 | 1.2 | 22 | 18 | 0.91 | 7.6 |
| IaFOC_rh | 0.52 | 0.28 | 6.4 | 3.7 | 12 | 5 | 2.8 | 1.1 | 0.66 | 3.3 | 0.42 | 1.8 | 1.5 | 0.38 | 0.49 | 17 | 8.3 | 0.8 | 4.2 |
| MTC_rh | 0.77 | 1.6 | 7.4 | 4.7 | 11 | 0.66 | 5.4 | 0.59 | 1 | 11 | 0.77 | 1.3 | 3.2 | 0.72 | 0.76 | 14 | 9.1 | 0.33 | 6.5 |
| LTC_rh | 0.37 | 1.2 | 9.6 | 4 | 12 | 1.3 | 4.2 | 0.47 | 0.28 | 9.7 | 0.59 | 3.3 | 3.2 | 0.54 | 1.9 | 20 | 18 | 0.31 | 7.6 |
| TJ_rh | 0.24 | 1.3 | 13 | 0.76 | 21 | 3.3 | 1 | 0.7 | 0.12 | 14 | 1.2 | 2.5 | 1.5 | 0.2 | 6.5 | 14 | 29 | 0.12 | 16 |
| SPC_rh | 0.6 | 1.1 | 7 | 3.7 | 17 | 12 | 2.6 | 0.66 | 0.99 | 15 | 3.3 | 6.6 | 1.4 | 0.023 | 10 | 18 | 44 | 0.2 | 17 |
| IPC_rh | 0.34 | 0.78 | 10 | 1.9 | 22 | 12 | 1.3 | 0.61 | 0.84 | 17 | 3.1 | 4.5 | 1.9 | 0.12 | 15 | 18 | 43 | 0.14 | 19 |
| PCC_rh | 1.8 | 1.2 | 10 | 8.5 | 24 | 8 | 2 | 0.7 | 1 | 16 | 2.5 | 9.5 | 1 | 0.15 | 14 | 19 | 28 | 0.61 | 22 |
| ACaMPC_rh | 0.36 | 0.66 | 2 | 10 | 10 | 3.6 | 3.4 | 0.4 | 0.24 | 4.1 | 1.4 | 1.7 | 0.91 | 0.052 | 1.4 | 6.5 | 2.2 | 0.37 | 0.74 |
| IFC_rh | 0.13 | 0.18 | 3.5 | 2.9 | 8.6 | 6.6 | 0.51 | 0.58 | 0.47 | 1.2 | 0.25 | 1.5 | 0.33 | 0.063 | 0.093 | 6.9 | 2.1 | 0.35 | 1.4 |
| DPC_rh | 0.2 | 0.41 | 2 | 5.1 | 6.2 | 4.9 | 1.4 | 0.43 | 0.57 | 3.4 | 0.96 | 1.5 | 0.47 | 0.032 | 0.42 | 4.5 | 2.6 | 0.39 | 0.64 |

Table T5: **EPR of HC session 2 across brain regions.**
